# Bi-allelic variants in *TBC1D8* result in non-obstructive azoospermia in both humans and mice

**DOI:** 10.1101/2025.10.31.25337980

**Authors:** Zihao Hu, Jun Zeng, Min Wen, Jing Zhang, Yongzhen Li, Jing Zhao, Jianlin Chen, Hongmei Xiao, Hualin Huang

**Affiliations:** Reproductive Medicine Center, Department of Obstetrics and Gynecology, the Second Xiangya Hospital, Central South University, Changsha, Hunan, China; Institute of Reproductive & Stem Cell Engineering, School of Basic Medical Science, Central South University, Changsha, Hunan, China; Hainan Provincial Key Laboratory for Human Reproductive Medicine and Genetic Research, Hainan Provincial Clinical Research Center for Thalassemia, Key Laboratory of Reproductive Health Diseases Research and Translation (Hainan Medical University), Ministry of Education the First Affiliated Hospital of Hainan Medical university, Hainan Medical University, Haikou Hainan, China; Department of Reproductive Medicine Center, The Third Xiangya Hospital, Central South University, Changsha, Hunan, China; Department of Pediatrics, the Third Xiangya Hospital, Central South University, Changsha, Hunan, China; Pediatric department, the Second Xiangya Hospital, Central South University, Changsha, Hunan, China; Reproductive Medicine Center, Xiangya Hospital, Central South University, Changsha, Hunan, China

**Keywords:** Infertility, Azoospermia, *TBC1D8*, RAB8A, Autophagy, Apoptosis

## Abstract

Non-obstructive azoospermia (NOA) is a severe form of male infertility. Proteins from the Tre2-Bub2-Cdc16 (TBC) family emerge as key contributors to spermatogenesis, including TBC1D20, TBC1D21, and TBC1D25. Nonetheless, the specific role of TBC1D8 in male male fertility remains unclear. Our study aims to elucidate the relationship between TBC1D8 and male infertility in humans and mice. Male *Tbc1d8*^-/-^ mice were completely infertile, producing no offspring. They exhibited spermiogenesis defects from steps 9 to 16, accompanied by pronounced acrosomal abnormalities during sperm maturation, ultimately preventing the formation of functional spermatozoa. Mechanistically, TBC1D8 deficiency impaired autophagic flux and enhanced apoptosis in the seminiferous epithelium through disrupted coupling with RAB8A, culminating in azoospermia and infertility. Consistent with the murine phenotype, whole-exome sequencing (WES) identified compound heterozygous missense variants in *TBC1D8* (NM_001102426.3) among three patients from unrelated Chinese Han families diagnosed with NOA or cryptozoospermia: Patient 1: c.3322A>G and c.1747C>T; Patient 2: c.3322A>G and c.2566A>G; Patient 3: c.845C>T and c.2525C>T. Histological examination of patient testicular biopsies revealed severely disrupted spermatogenic architecture and markedly reduced TBC1D8 expression, suggesting a loss-of- function effect. Together, our findings establish TBC1D8 as a previously unrecognized regulator of spermatogenesis, where its deficiency leads to NOA through impaired autophagy and increased apoptosis..

## Introduction

Globally, approximately 10-20% of couples face infertility(1). Calculated data show that male factors contribute to infertility, ranging from 20% to 70%, with the prevalence of male infertility spanning between 2.5% and 12%(2). Male infertility can result from abnormalities in sperm quantity or quality, of with non-obstructive azoospermia (NOA) representing the most severe manifestation due to impaired spermatogenesis. Histological manifestations of the testis in NOA encompass Sertoli-Cell-Only Syndrome (SCOS), Maturation Arrest (MA), and hypospermatogenesis(3). Genetic factors explain approximately 20-25% of azoospermia cases, including karyotype abnormalities, Azoospermia Factor (AZF) microdeletions and some monogenic causes(3–5). The etiology of over 70% of NOA cases remains unknown, termed idiopathic NOA, warranting further investigation.

The TBC protein family constitutes a highly evolutionarily conserved superfamily, characterized by the Tre2-Bub2-Cdc16 (TBC) domain. In humans, at least 40 proteins contain this TBC domain. Previous reports have linked defects in *TBC1D20* (OMIM: 615663), *TBC1D21* (OMIM: 620387), and *TBC1D25* genes to male sterility(6–8). *TBC1D20* significantly contributes to acrosome formation in sperm and the maintenance of the blood- testis barrier integrity(9–11). Similarly, *TBC1D21* is involved in upholding the structural integrity of the mitochondrial sheath and tail axoneme in sperm(7, 12). *TBC1D25* variant has been reported in oligozoospermia patients(8). Overall, TBC family proteins emerge as key contributors to spermatogenesis, exerting a substantial impact on male fertility.

TBC1D8, a member of the TBC family proteins, is less well-studied. It has been reported to facilitate cell proliferation and metabolic regulation in cancers, associated with a poorer prognosis in patients (13, 14). Decreased methylation of the *TBC1D8* gene promoter CpG islands may predict the risk for osteoporosis(15). Nonetheless, the specific role of *TBC1D8* in male fertility remains ambiguous at present.

Here, we demonstrate that *TBC1D8* is indispensable for spermatogenesis. Male *Tbc1d8*^-/-^ mice were completely sterile, exhibiting spermiogenesis defects during the late stages of germ cell differentiation and severe acrosomal abnormalities. Mechanistically, TBC1D8 deficiency disrupted autophagic flux and triggered extensive germ cell apoptosis within the seminiferous epithelium, ultimately leading to azoospermia. Consistent with the murine phenotype, we identified compound heterozygous missense variants of *TBC1D8* (NM_001102426.3) in three infertile male patients from unrelated Chinese Han families (Patient 1: c.3322A>G and c.1747C>T; Patient 2: c.3322A>G and c.2566A>G; Patient 3: c.845C>T and c.2525C>T). Clinical and histological analyses of patient samples revealed impaired spermatogenic architecture and markedly reduced TBC1D8 expression, supporting a loss-of-function effect. Our findings establish *TBC1D8* as a novel genetic determinant of NOA and suggest that its screening may facilitate diagnosis and provide a rationale for future therapeutic interventions.

## Materials and methods

### Generation of *Tbc1d8* knockout mice

CRISPR/Cas9 was used to generate *Tbc1d8* (GenBank: NM_018775.4) knockout (KO) mice. The guide RNA (gRNA) sequences and genotyping primers are detailed in the Supplementary Materials (Supplementary Fig. S1 and Table S2 and 3). The mice were housed under specific- pathogen-free conditions within the Laboratory Animal Center of Hunan University of Chinese Medicine. The Animal Ethics Committee of Hunan University of Traditional Chinese Medicine approved the ethical review of animal experimentation.

### Fertility test

Each 8-week-old male mouse was caged with two wild-type female mice at the same age for 3Lmonths and all the females were monitored for pregnancy. The total numbers of pups were recorded for all the litters.

### Human subjects

All three patients underwent comprehensive clinical assessments. Physical examination and detailed medical history revealed no congenital abnormalities, such as undescended testicles, hypospadias, or hypoplastic seminal vesicles. Additionally, no incidental findings including varicocele, urinary tract infection, or orchitis were observed. None of the patients had a history of exposure to gonadotoxic agents, such as radiation or chemotherapy. All patients also underwent comprehensive andrological evaluation, including physical examination and scrotal Doppler ultrasound, independently performed by two experienced andrologists. The presence or absence of varicocele was assessed, and no evidence of varicocele was detected in any of the participants. Semen analysis was conducted following the World Health Organization 6th guidelines(16). Moreover, serum sex hormone testing (FSH, LH, PRL and testosterone), chromosome karyotype analysis, and azoospermia factor microdeletion analysis were performed. Tissue samples were collected and processed under the same conditions for both NOA and obstructive azoospermia (OA) patients to ensure consistency and comparability. Testicular tissues from patient with OA were used as a normal control (NC). Testicular Sperm Aspiration (TESA) was initially performed for diagnostic purposes, followed by microsurgical Testicular Sperm Extraction (micro-TESE) for sperm retrieval intended for intracytoplasmic sperm injection (ICSI). Overall, the patients were clinically unremarkable aside from their respective diagnoses of NOA or cryptozoospermia. Participants granted informed consent for the collection of semen, testis tissue and peripheral blood. This study was approved by the Ethics Committee of the Second Xiangya Hospital, Central South University.

### Exome sequencing and *in silico* bioinformatics analysis

Peripheral blood samples were collected from patients without chromosomal abnormalities, azoospermia factor microdeletions, or other evident causes. Exome sequencing was conducted using the Agilent SureSelect Exome Enrichment Kit V6 and the Illumina HiSeq 3000 platform. According to ExAC 0.3.1 and gnomAD v4.1.0, variants with allele frequencies over 5% were excluded. Nonsense, frameshift, splice, and missense variants were initially selected for analysis using tools such as SIFT, MutationTaster, PROVEAN, CADD, and fathmm-MKL(Supplementary Table S1).Sanger sequencing was employed to validate variant presence within family members. PCR and Sanger sequencing were performed to verify variants in the probands and their parents. The primers are listed in Supplementary Materials(Supplementary Table S2.

To assess the impact of candidate variants, changes in protein structure were predicted using Alphafold2 based on the J3KQ40 PDB template(17, 18). The effects of missense variants on three-dimensional structures were visualized using PyMOL v1.7.4.

### Haematoxylin–eosin (HE) staining and periodic acid–Schiff (PAS) staining

The testicular tissues were fixed in 4% paraformaldehyde for 24 hours, followed by embeddeding in paraffin, and subsequently. The tissues were then sectioned to a thickness of 4 mm for furthersubsequent examination. The sections were stained using hematoxylin, Eosin, and Schiff’s solution (Abiowell, AWI0020a/bioss, S0127) according to the manufacture instructions. Images of the stained sections were captured using a scanner (Kfbio, KF-PRO- 005-EX).

### Immunohistochemistry (IHC) and immunofluorescence (IF)

The sections of Mouse or human testes were deparaffinized in xylene, dehydrated in a decreasing alcohol gradient, and underwent antigen retrieval by heating in sodium citrate buffer. Following blocking with 5% BSA, the sections were incubated with the primary antibody overnight at 4°C. Subsequently, the sections were incubated with an HRP- conjugated secondary antibody and developed into color using DAB (ZSGB-BIO, ZLI-9018). Nuclear staining was achieved with Hematoxylin before capturing images. In immunofluorescence experiments, sections were incubated with Alexa Fluor 488 or Alexa Fluor 555-coupled secondary antibodies and counterstained with antifade mounting medium with DAPI (Beyotime, P0131) before observation under a fluorescence microscope. Details of the antibodies used are provided in the Supplementary Materials(Supplementary Table S4).

### Co-immunoprecipitation (co-IP) and Western blot (WB)

Eight-week-old male mice were randomly selected. Following euthanization, the spleens were extracted and lysed using RIPA to isolate proteins. Protein lysates were then incubated with an anti-TBC1D8 antibody for 24 hours with shaking at 4L. Subsequently, Protein A+G Agarose Beads (Beyotime, P2108) were added and shaken for 4 hours at room temperature to capture the proteins that interacteding with TBC1D8.

The proteins were separated by the SDS-PAGE and then transferred to thea PVDF membrane. The membrane was blocked with a protein-blocking solution (Boster, AR0041), and primary antibodies were incubated on it overnight at 4°C. After beFollowing a washed with TBST, the membranes were incubated with an HRP-conjugated secondary antibody. The image capture was accomplished by utilizwas performed using a chemiluminescence detection system (Bio- Rad, ChemiDoc XRS+ System). Details of the antibodies utilizsed weare listed in the Supplementary Materials (Supplementary Table S4).

### Ethics Statement

The animal study was reviewed and approved by the Animal Ethics Committee of Hunan University of Traditional Chinese Medicine approved the ethical review of animal experimentation. All methods were performed in accordance with the relevant guidelines and regulations. For study patients,this study was approved by the Ethics Committee of the Second Xiangya Hospital, Central South University. All the studies were carried out in accordance with the Declaration of Helsinki and approved by the institutional ethics review board.

### Statistical analysis

All analyses were performed following a minimum of 3 (n≥3) independent experiments. Unpaired, two-tailed Student’s t test or Welch’s t test were employed for statistical analysis. A p-value less than 0.05 is deemed statistically significant.

## Results

### TBC1D8 deficiency caused spermatogenesis defects and male infertility in mice

To investigate the in vivo function of TBC1D8 in spermatogenesis, we employed a mouse model, given the 91.97% similarity between the amino acid sequence of mouse and human TBC1D8 proteins (Supplementary Fig. S2). To elucidate the pathogenic mechanisms associated with TBC1D8 deficiency in NOA, we used CRISPR/Cas9 technology to generate *Tbc1d8* knockout (*Tbc1d8^-/-^*) mice by deleting exon 3 to exon 14 (Supplementary Fig. S1). The body weight and testicular volume of *Tbc1d8*^-/-^ mice didn’t decrease significantly compared to *Tbc1d8^+/+^* mice (Fig. 1a and b). Effective knockout was confirmed through WB and IHC assessments (Fig. 1c-f). Each 8-week-old male mouse was caged with two wild-type female mice at the same age for 3 months. Compared with the *Tbc1d8^+/+^* mice, *Tbc1d8*^-/-^ mice were totally infertile (Table 1).

**Figure 1.**
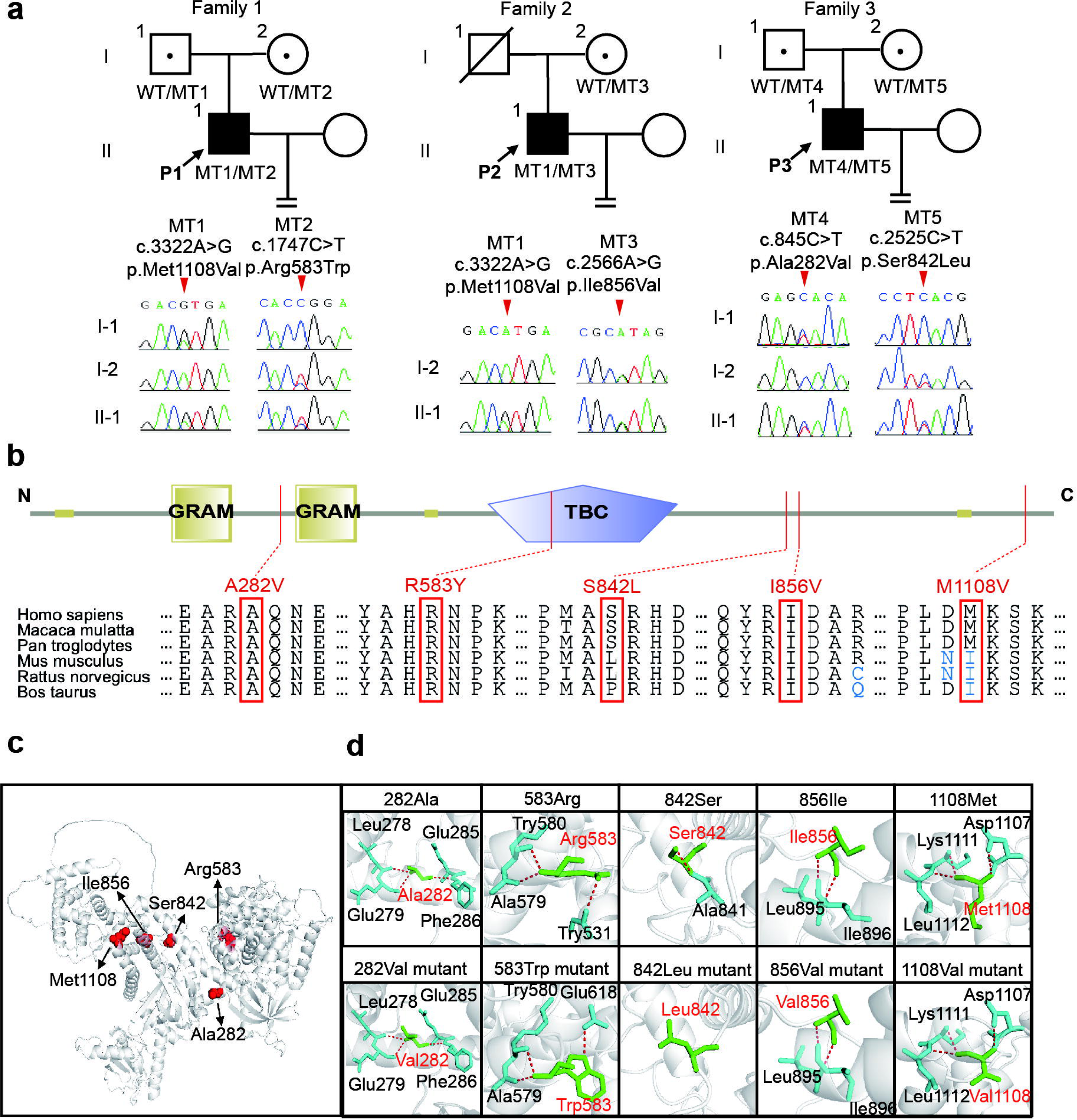
Generation of *Tbc1d8* knockout mice. (A) The physical and testicular development of *Tbc1d8^+/+^* and *Tbc1d8^-/-^* male adult mice are normal. (B) Testis/body weight of *Tbc1d8^+/+^* and *Tbc1d8^-/-^* male adult mice. Data are represented as individual values with mean ± SD. n = 3 biological replicates. Unpaired 2-sided t test. (C) WB analysis of TBC1D8 and DDX4 expression in the testis from *Tbc1d8^+/+^* and *Tbc1d8^-/-^* male mice. (D) Relative protein levels of TBC1D8 and DDX4 in testes from *Tbc1d8^+/+^* and *Tbc1d8^-/-^*male mice. Relative amounts of proteins were calculated after normalization to the GAPDH protein level. Data are represented as individual values with mean ± SD. n = 3 biological replicates. The experiments were repeated three times. Unpaired 2-sided t test. (E) Immunohistochemistry analysis of TBC1D8 expression in the testis from *Tbc1d8^+/+^* and *Tbc1d8^-/-^* male mice. Scale bars denote 100μm and 10μm. (F) Intensity of TBC1D8 in the testis of *Tbc1d8^+/+^* and *Tbc1d8^-/-^* male mice. Data are represented as individual values with mean ± SD. n = 3 biological replicates. The experiments were repeated three times. Unpaired 2-sided t-test. (G) Immunofluorescence analysis of DDX4 expression in the testis from *Tbc1d8^+/+^* and *Tbc1d8^-/-^* male mice. Scale bars denote 50μm and 10μm. (H) Fluorescence intensity of DDX4 in the testis of *Tbc1d8^+/+^* and *Tbc1d8^-/-^* male mice. Data are represented as individual values with mean ± SD. n = 3 biological replicates. The experiments were repeated three times. Unpaired 2-sided t-test. (I) HE-stained histological sections of testes from *Tbc1d8^+/+^*and *Tbc1d8^-/-^* male mice. The asterisk represents apoptotic spermatocyte. Yellow triangle represents multinucleated cells (symblasts). Blue arrow represents vacuolated seminiferous tubules. White arrow represents detachment of sperm cells. Blue triangle represents cytoplasm resembling residual bodies. Red arrow represents ill-shaped nuclei. Scale bars denote 50μm. (J) HE-stained histological sections of the epididymis from *Tbc1d8^+/+^* and *Tbc1d8^-/-^* adult male mice. Scale bars denote 100μm. \**P* < 0.05; \*\**P* < 0.01; \*\*\**P* < 0.001; \*\*\*\**P* < 0.0001.

**Table 1.** Male mouse fertility assay.

| Genotype | Mating period | Nunber of fertile male mice test | Total pups |
| --- | --- | --- | --- |
| *Tbc1d8<sup>+/+</sup>* | 3 months | 3/3 | 78 |
| *Tbc1d8<sup>-/-</sup>* | 3 months | 0/3 | 0* |

\**P* < 0.05

TBC1D8 exhibited expression in various stages of germ cells but not in the Sertoli cells, particularly in spermatocytes and spermatids, displaying strong intensity in the testes of *Tbc1d8^+/+^* mice (Fig. 1e and f). However, histological analysis revealed a notable reduction in the number of germ cell in seminiferous tubules of *Tbc1d8^-/-^*mice by the decline of DDX4 expression, the marker of germ cells (Fig. 1c and d). IF staining correspondingly revealed a notable reduction in the number of DDX4-positive cells within the seminiferous tubules of *Tbc1d8^-/-^* mice compared with *Tbc1d8^+/+^* mice (Fig. 1g-h).

To clarify the phenotype of spermatogenic retardation, HE staining was performed on the testis and epididymis of mice. *Tbc1d8^-/-^*mice exhibit incomplete MA in the seminiferous epithelium, characterized by abnormal spermatocytes and lack of mature spermatids, leading to NOA and infertility. On the one hand, abnormalities in spermatogenesis in *Tbc1d8^-/-^* mice were characterized by the absence of elongated and mature sperm, with the accumulation of cytoplasm resembling residual bodies, multinucleated cells, ill-shaped nuclei, and the detachment of round spermatids (Fig. 1i). On the other hand, vacuolated seminiferous tubules were observed in the testis of *Tbc1d8^-/-^* mice, accompanied by the presence of apoptotic spermatocyte (Fig. 1i). Additionally, the majority of epididymal tubules in *Tbc1d8^-/-^* mice lacked spermatozoa, displaying only a few round spermatids or multinucleated cells, and residual bodies in the caput epididymis and cauda epididymis (Fig. 1j).

Round spermatids were seen in the seminiferous tubules, but elongated spermatids were deficient, suggesting impaired spermiogenesis in *Tbc1d8^-/-^*mice. PAS staining showed the lack of elongated spermatids from stages I to VIII, with an abundance of accumulated residual bodies (Fig. 2a). Further observation of head shaping between *Tbc1d8^+/+^* and *Tbc1d8^-/-^* mice was compared. During steps 1 to 8, there were no obvious abnormalities in the formation of the acrosome of round spermatids, with the acrosome continuing to expand along the nuclear membrane as it deforms (Fig. 2b). Starting from step 9, the round spermatids began to elongate. The spermatids of *Tbc1d8^-/-^* mice began to have morphological abnormalities from step 12, manifested by the failure of acrosome extension, pyknosis of the nucleus, and ultimately the inability to produce mature spermatozoa (Fig. 2b). Statistical analysis of the number of normal spermatids showed a significant decrease from steps 9 to 16 (stage I to VIII and stage XI to XII) (Fig. 2b-c). In summary, the deletion of *Tbc1d8* in male mice resulted in impaired spermiogenesis from step 9 to 16, leading to the inability to produce mature spermatozoa. In brief, in the Golgi and Cap phases *Tbc1d8^-/-^* acrosomal structures appear normal, but abnormalities become apparent in the Maturation phase.

**Figure 2.**
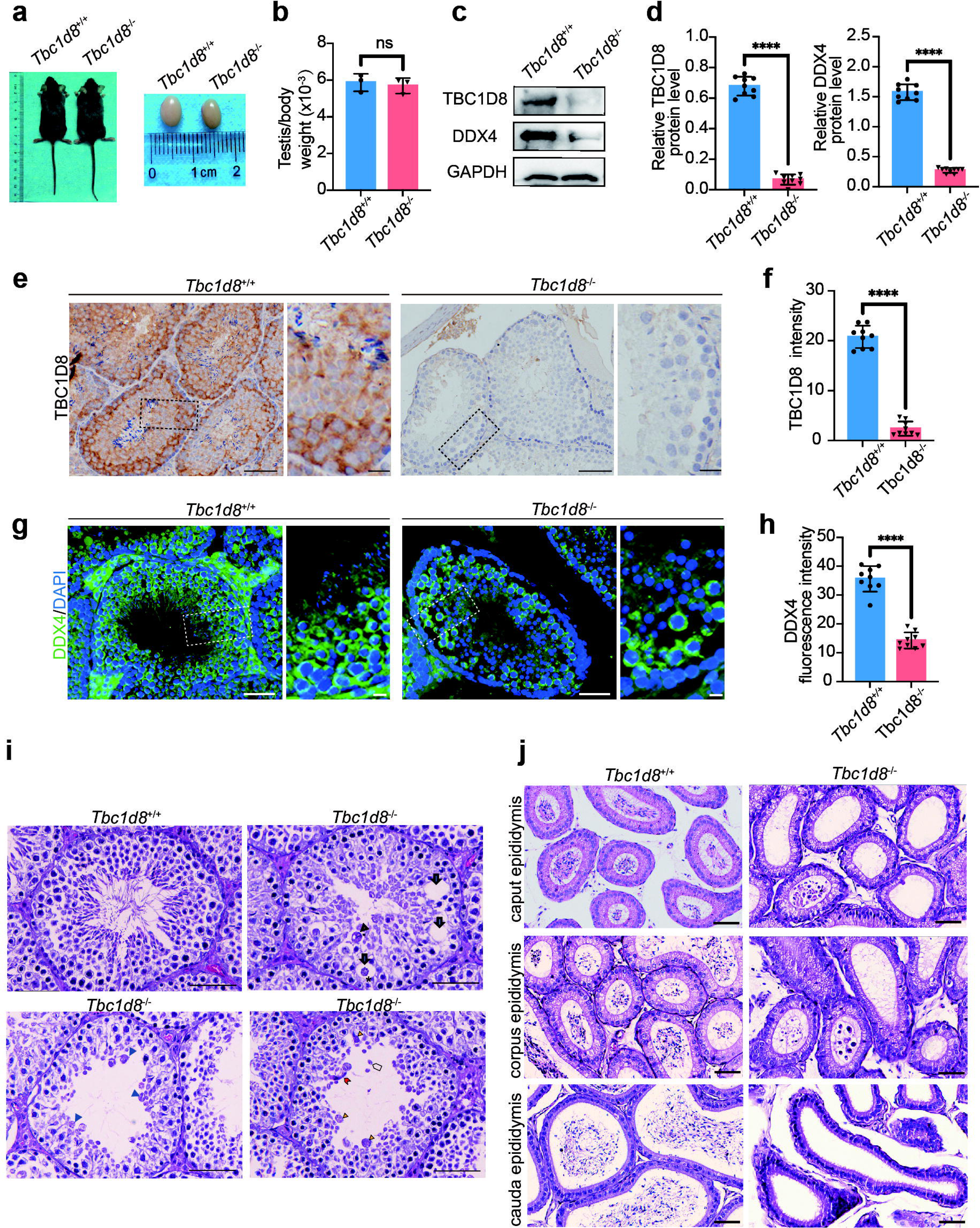
*Tbc1d8^-^*^/-^ male mice displayed spermatogenesis defects. (A) PAS staining of testes from *Tbc1d8*^+/+^ and *Tbc1d8*^-/-^ male mice. Cycle of the seminiferous epithelium are shown from stage I to stage XII. M, metaphase of meiosis I spermatocyte; rST, round spermatid; eST, elongating spermatids. Scale bars denote 20μm. (B) PAS staining of spermatids at different developmental stages is shown from steps 1 to step 16. Scale bars denote 1μm. (C) The graphical representation illustrates the spermatid number at different developmental steps (steps 1 to 16) using box-and-whisker plots. The key components of the plots are defined as follows: Bounds of the Boxes represent the interquartile range. Lines within the Boxes indicate the median. Whiskers extend from the boxes to encapsulate the full range of data. n = 3 biological replicates. Unpaired 2-sided t-test. \**P* < 0.05; \*\**P* < 0.01; \*\*\**P* < 0.001; \*\*\*\**P* < 0.0001.

### RAB8A interacted with TBC1D8 and decreased in *Tbc1d8^-/-^* mice

As TBC/RAB-GAP proteins play a crucial role in vesicle transport, we employed co-IP combined with mass spectrometry to identify RAB8A as an interacting partner of TBC1D8 (Fig.3a and b). IF results demonstrated the co-localization of TBC1D8 with RAB8A in spermatocytes and elongated spermatids (Fig. 3e). Furthermore, the expression of RAB8A showed a proportional decrease with TBC1D8 levels in the testis, with minimal expression observed in *Tbc1d8^-/-^*mice compared to *Tbc1d8^+/+^* mice (Fig. 3c-e). These findings suggest that TBC1D8 may regulate spermatogenesis in spermatocytes and spermiogenesis in spermatozoa through its interaction with RAB8A.

**Figure 3.**
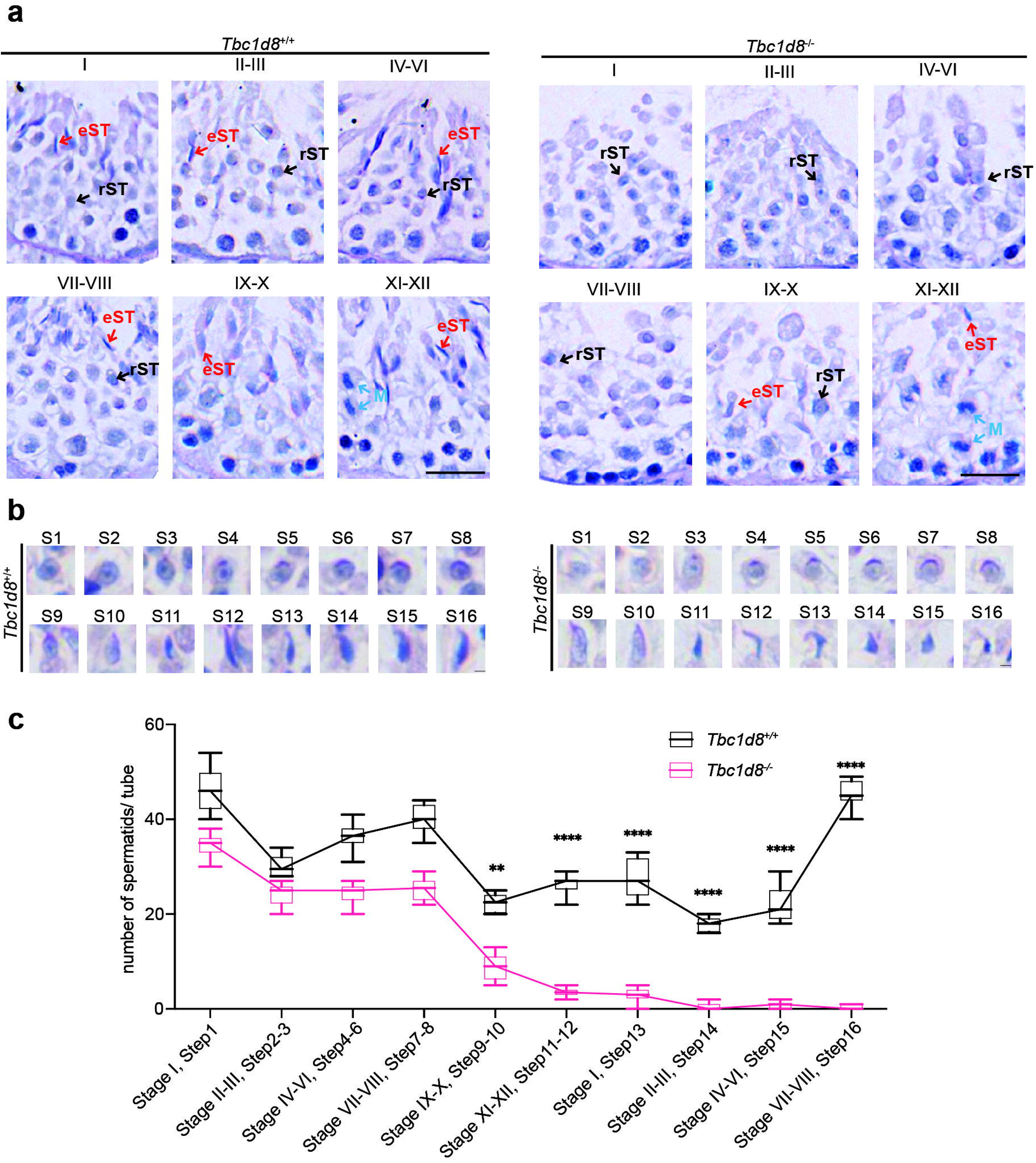
RAB8A interacts with TBC1D8 and decreased in *Tbc1d8^-/-^* mice. (A) Immunoprecipitation-Mass Spectrometry analysis using anti-TBC1D8 antibody revealed the unique peptide of RAB8A. (B) Co-IP and WB analysis confirmed the interaction between TBC1D8 and RAB8A. (C) WB analysis for the expression of RAB8A in the testis of *Tbc1d8^+/+^* and *Tbc1d8^-/-^* adult male mice. (D) Relative protein levels of RAB8A in the testes of *Tbc1d8^+/+^* and *Tbc1d8^-/-^* male mice. Relative amounts of proteins were calculated after normalization to the GAPDH protein level. Data are represented as individual values with mean ± SD. n = 3 biological replicates. The experiments were repeated 3 times. Unpaired 2-sided t test. (E) Immunofluorescence analysis for colocalization of TBC1D8 and RAB8A in the testes of *Tbc1d8^+/+^*and *Tbc1d8^-/-^* mice. Scale bars denote 50μm.\**P* < 0.05; \*\**P* < 0.01; \*\*\**P* < 0.001; \*\*\*\**P* < 0.0001.

### TBC1D8 deletion suppressed the autophagosome maturation

Autophagy is critical for the spermiogenesis process. Previous research has reported that TBC1D20 mediates autophagy as a key regulator of autophagosome maturation through RAB1B(11). Compared to *Tbc1d8^+/+^* mice, testicular tissues of *Tbc1d8^-/-^*mice showed a notable increase in ATG7 accumulation, LC3II conversion, and P62 accumulation(Fig. 4a and b). ATG7, a protein that regulates autophagosome formation, was primarily located within the cytoplasm resembling residual bodies that abnormally aggregate in the *Tbc1d8^-/-^* mice tubules (Fig. 4c). This could potentially exacerbate the buildup of autophagosomes within the seminiferous epithelium. Notably, ATG3 and ATG5 did not exhibit this accumulation (Fig. 4a and b). To investigate the impact of the concurrent downregulation of TBC1D8 and RAB8A on autophagic flux, we utilized IF staining to detect the autophagy markers LC3 and P62 in mouse testis. In *Tbc1d8^+/+^* seminiferous tubules, we observed crescentic staining of LC3 and P62, indicating their presence at the acrosomal location of acrosomes (Fig. 4c, 5b and c). However, in *Tbc1d8^-/-^* mice, significant and diffuse accumulation of LC3 and P62 was observed throughout the germinal epithelium, suggesting impaired lysosomal degradation (Fig. 4c and d, 5b and c), resembling observations from a TBC1D8-deficient Drosophila model(19). Peanut agglutinin (PNA) staining showed a lack of mature crescent-shaped acrosomes within the elongated spermatozoa of *Tbc1d8^-/-^*mice (Fig. 5a-c). These findings suggest that TBC1D8 deficiency might lead to aberrant interaction with RAB8A and increase in ATG7, impairing autophagosome maturation and blocking autophagic flux, consequently resulting in delayed spermiogenesis.

**Figure 4.**
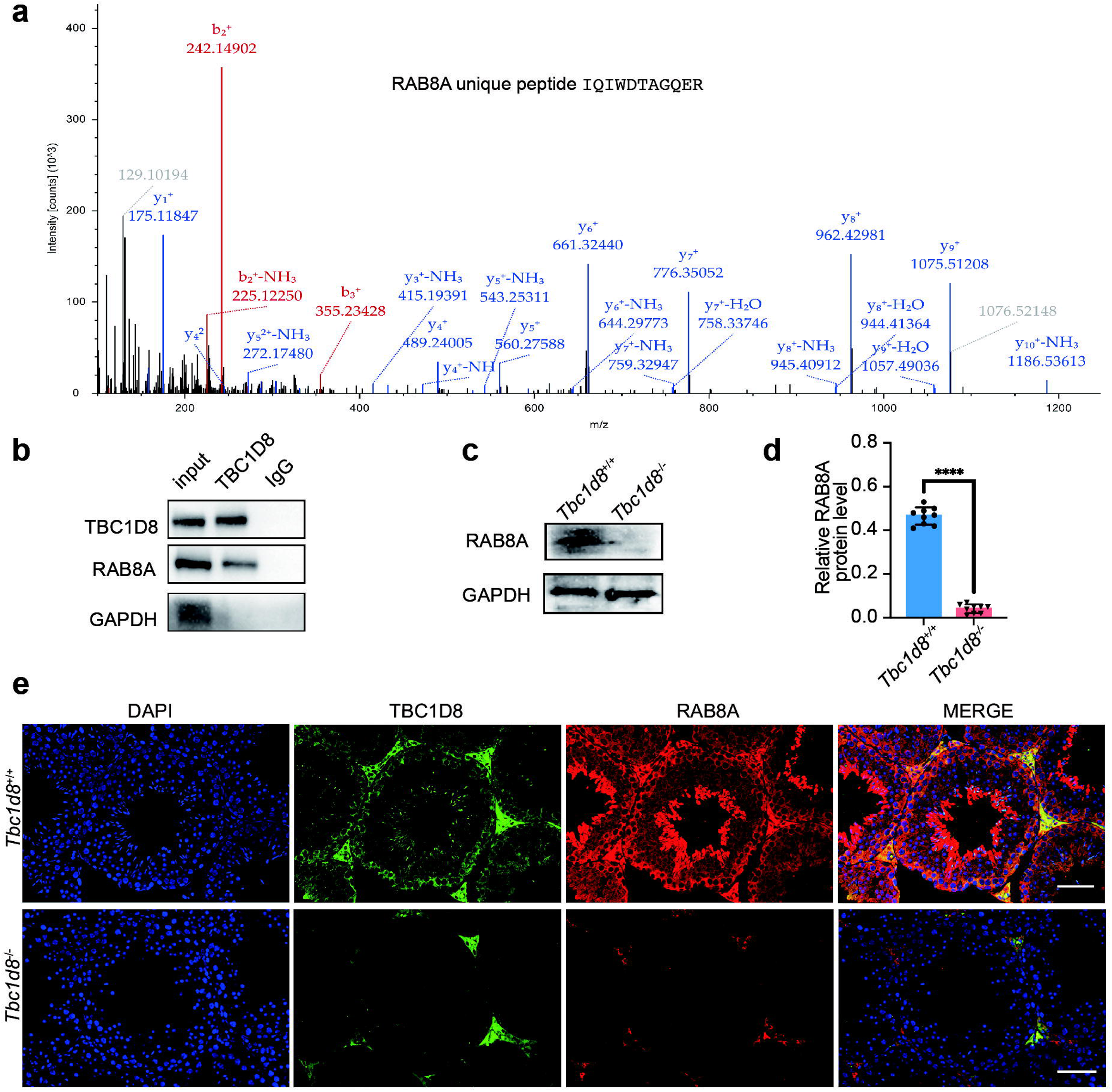
TBC1D8 deletion suppressed the autophagosome maturation. (A) WB analysis for autophagy signaling molecules ATG3, ATG5, ATG7, P62, and LC3 in the testis of *Tbc1d8^+/+^* and *Tbc1d8^-/-^* adult male mice. (B) Relative protein levels of ATG3, ATG5, ATG7, P62, and LC3 in the testes of *Tbc1d8^+/+^* and *Tbc1d8^-/-^* male mice. Relative amounts of proteins were calculated after normalization to the α-tubulin protein level. Data are represented as individual values with mean ± SD. n = 3 biological replicates. The experiments were repeated 3 times. Unpaired 2-sided t test. (C) Immunofluorescence analysis for ATG7, LC3, and P62 expression in the testis from *Tbc1d8^+/+^* and *Tbc1d8^-/-^* adult mice. Scale bars denote 50μm and 10μm. (D) Fluorescence intensity of ATG7, LC3, and P62 in the testis of *Tbc1d8^+/+^* and *Tbc1d8^-/-^*male mice. Data are represented as individual values with mean ± SD. n = 3 biological replicates. The experiments were repeated 3 times. Unpaired 2-sided t test. \**P* < 0.05; \*\**P* < 0.01; \*\*\**P* < 0.001; \*\*\*\**P* < 0.0001.

**Figure 5.**
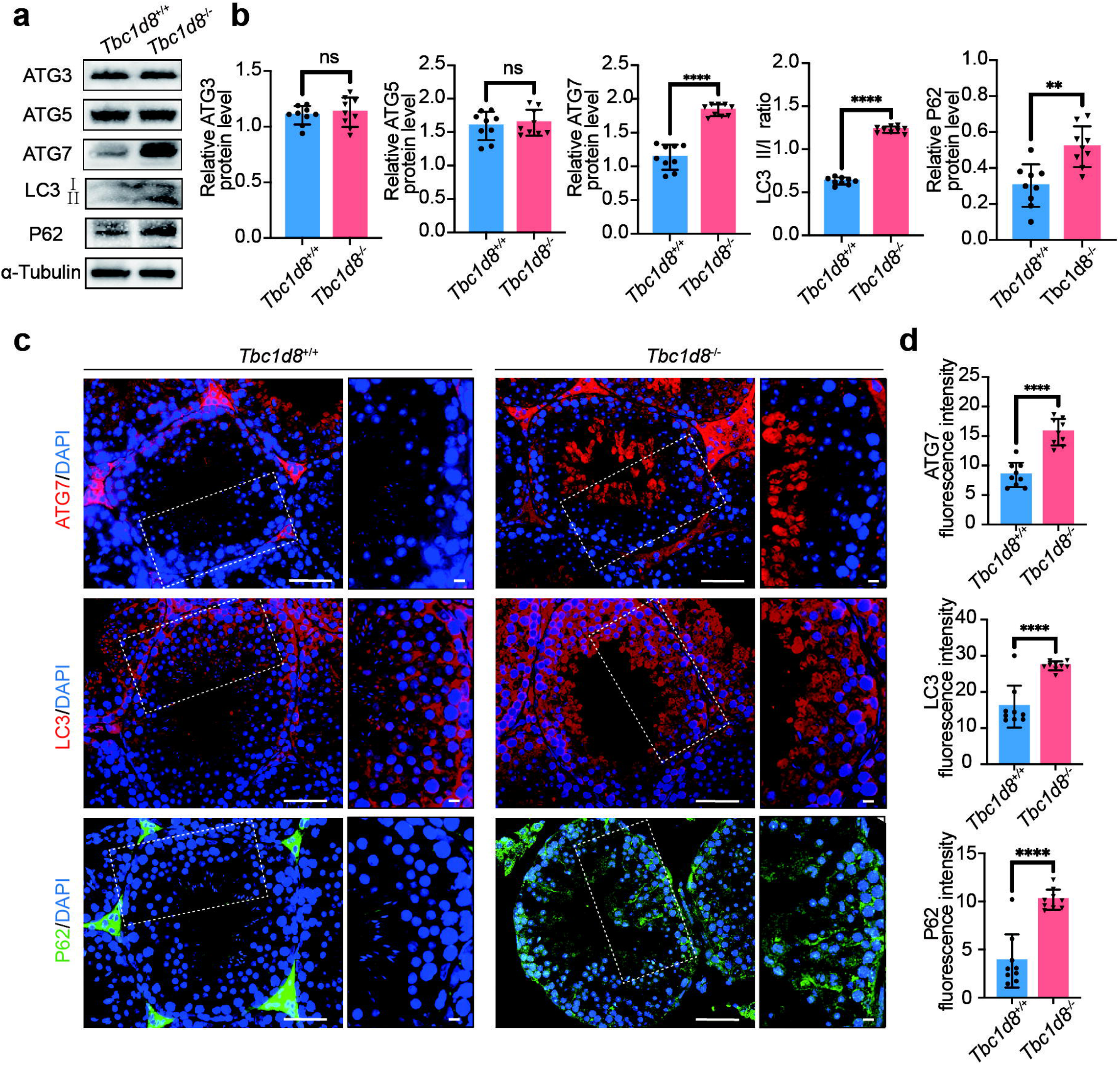
Autophagy defects lead to impaired acrosome formation (A) FITC-labeled PNA staining for acrosome formation analysis. Scale bars denote 20μm. (B) Immunofluorescence analysis for LC3 and PNA colocalization in the testis from *Tbc1d8^+/+^* and *Tbc1d8^-/-^* adult mice. Scale bars denote 20μm. (C) Immunofluorescence analysis for P62 and PNA colocalization in the testis from *Tbc1d8^+/+^* and *Tbc1d8^-/-^* adult mice. Scale bars denote 20μm.

### TBC1D8 deprivation induced abnormal apoptosis in seminiferous tubules

Previous studies have indicated that RAB8A deficiency leads to apoptosis in other diseases (20). To clarify the reason for vacuolization of testicular spermatocytes in *Tbc1d8*^-/-^ mice, we examined indicators related to apoptosis. Compared with *Tbc1d8*^+/+^ mice, the testes of *Tbc1d8*^-/-^ mice exhibited increased expression of the apoptotic protein BAX, down-regulation of the anti-apoptotic protein BCL2, and elevated levels of active Caspase-3 (Fig. 6a and b). IHC and IF results showed that pro-apoptotic proteins accumulated in spermatocytes while the expression of anti-apoptotic proteins decreased, leading to apoptosis of spermatocytes (Fig. 6c-f). The TUNEL assay revealed an increased average TUNEL-positive cells in the testes of *Tbc1d8^-/-^* mice (Fig. 6g and h). In addition, apoptosis also occurred in some detached round spermatids and ill-shaped spermatozoa, as shown by TUNEL staining of the epididymis (Fig. 6g and i). Together, these findings confirmed that TBC1D8 deletion, coupled with reduced RAB8A expression, led to increased apoptosis in the male testes.

**Figure 6.**
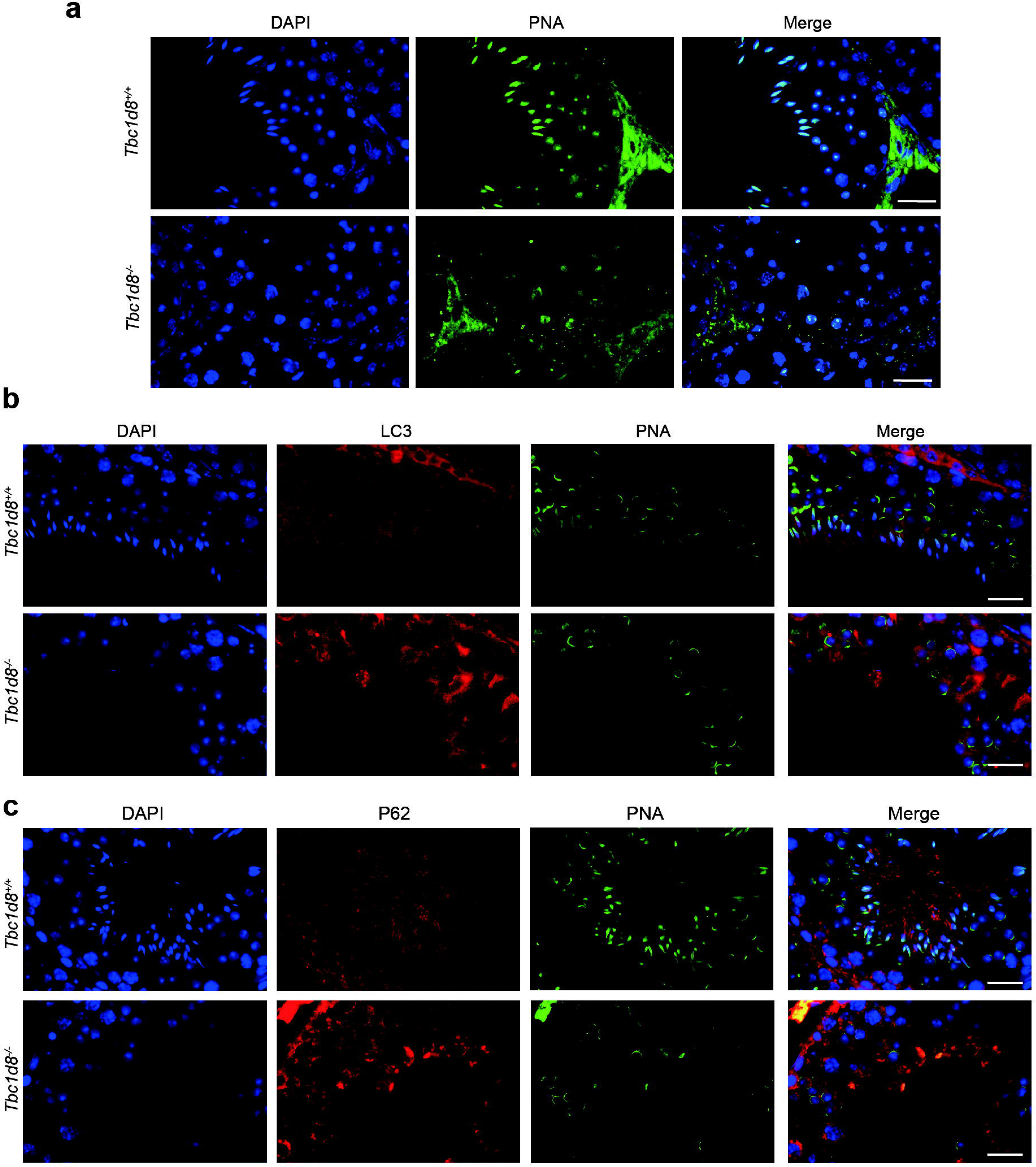
TBC1D8 deprivation caused abnormal apoptosis in seminiferous tubules. (A) WB analysis for apoptosis signaling molecules BAX, BCL2, and Cleaved-caspase3 in the testis of *Tbc1d8^+/+^* and *Tbc1d8^-/-^* adult male mice. (B) Relative protein levels of BAX, BCL2, and Cleaved-caspase3 in the testes of *Tbc1d8^+/+^* and *Tbc1d8^-/-^* male mice. Relative amounts of proteins were calculated after normalization to the α-tubulin protein level. Data are represented as individual values with mean ± SD. n = 3 biological replicates. The experiments were repeated 3 times. Unpaired 2-sided t test. (C) Immunohistochemistry analysis of BAX and BCL2 expression in the testis of *Tbc1d8^+/+^* and *Tbc1d8^-/-^* male mice. Scale bars denote 50μm and 10μm. (D) Intensity of BAX and BCL2 in the testis of *Tbc1d8^+/+^* and *Tbc1d8^-/-^* male mice. Data are represented as individual values with mean ± SD. n = 3 biological replicates. The experiments were repeated 3 times. Unpaired 2-sided t test. (E) Immunofluorescence analysis for Cleaved-caspase3 expression in the testis of *Tbc1d8^+/+^* and *Tbc1d8^-/-^* adult mice. Scale bars denote 50μm and 10μm. (F) Fluorescence intensity of Cleaved-caspase3 in the testis of *Tbc1d8^+/+^* and *Tbc1d8^-/-^* male mice. Data are represented as individual values with mean ± SD. n = 3 biological replicates. The experiments were repeated 3 times. Unpaired 2-sided t test. (G) TUNEL assay in the testis and epididymis of *Tbc1d8^+/+^* and *Tbc1d8^-/-^* adult mice. Scale bars denote 50μm. (H) Graph showing the average number of TUNEL positive cells in the testis of *Tbc1d8^+/+^* and *Tbc1d8^-/-^* male mice. The largest cross-section of each testis was used for staining and cell counting (nL=L3). Data are represented as individual values with mean ± SD. The experiments were repeated 3 times. Unpaired 2-sided t test. (I) Graph showing the percentage of TUNEL positive tube in the epididymis of *Tbc1d8^+/+^* and *Tbc1d8^-/-^* male mice. The largest cross-section of each epididymis was used for staining and cell counting (nL=L3). Data are represented as individual values with mean ± SD. The experiments were repeated 3 times. Unpaired 2-sided t test. \**P* < 0.05; \*\**P* < 0.01; \*\*\**P* < 0.001; \*\*\*\**P* < 0.0001.

### Identification of *TBC1D8* pathogenic variants in NOA patient

We conducted whole exome sequencing (WES) on the probands to elucidate the genetic cause. Since the parents of the probands were capable of producing offspring without a similar family history, our focus was on variants following an autosomal recessive inheritance pattern (Fig. 7a).

**Figure 7.**
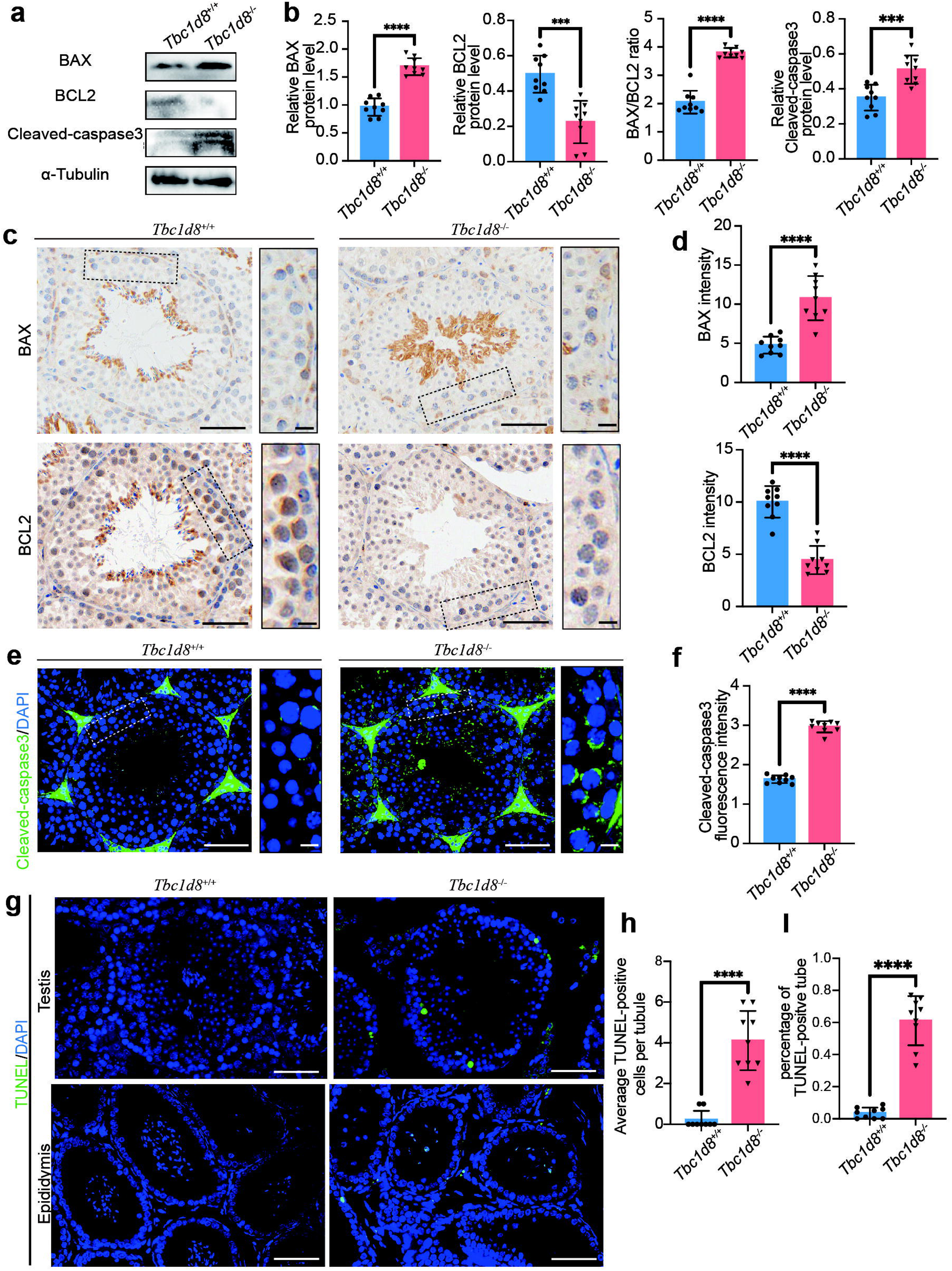
Identification of compound variants in *TBC1D8* from NOA-affected individuals. (A) Pedigrees of the 3 Chinese Han families affected by male infertility with Sanger sequencing validation for the compound heterozygous variants in *TBC1D8* (NM_001102426.3). Geometric symbols represent genders: squares denote males, circles denote females. Affected individuals are marked with black symbols, while unaffected ones are denoted by unfilled symbols. The obligate carrier is denoted with a dot-containing circle. Black-filled squares specifically highlight male individuals exhibiting NOA and cryptozoospermia. The position of the variant is indicated by red triangles. P1-3, patient 1-3. MT1–5, variants 1–5. (B) Schematic representation of TBC1D8 domains and the location of variant sites. GRAM, gucosyl transferases RAB-like GTPase activators, and myotubularins domain; TBC, Tre2- Bub2-Cdc16 domain. Amino acid conservation analysis of *TBC1D8* variants. The amino acids encoded by the variant sites are highlighted using a red box. (C) Protein structure and the location of variant sites. Variant positions are highlighted in red, and black arrows indicate the corresponding amino acids. (D) Changes in amino acids and hydrogen bonds after missense variant, analyzed by Alphafold2 and PyMol software. Hydrogen bonds are represented by red dotted lines.

Sanger sequencing was employed to validate the genetic evidence for *TBC1D8* in the probands. In total, 5 missense variants were identified in *TBC1D8* (NM_001102426.3) from three infertile male patients (Fig. 7a). In patient 1 (P1) and patient 2 (P2), both of them carried compound heterozygous variants, including the recurrent variant c.3322A>G (p.Met1108Val), along with c.1747C>T (p.Arg583Trp) and c.2566A>G (p.Ile856Val) respectively. The patient 3 (P3) exhibited compound heterozygous missense variants c.845C>T (p.Ala282Val) and c.2525C>T (p.Ser842Leu). The variants in P1 and P3 were inherited from the father and mother, respectively. In P2, one variant was confirmed to be maternally inherited, whereas the other variant may be paternally inherited or represent a de novo mutation (Fig. 7a). Furthermore, all identified *TBC1D8* variants exhibited extremely low frequencies in the ExAC Browser and gnomAD databases (Supplemetary Table S1). The amino acids encoded by the variant sites exhibited high conservation in primates (Fig. 7b). Pathogenic analysis suggested that these missense variants were likely to be deleterious or detrimental (Supplemetary Table S1). The amino acid variations were predicted to impact the protein structures and amino acid interactions using Alphafold2 (Fig. 7c-d). Specifically, p.Arg583Trp and p.Ser842Leu were identified as having the potential to alter the hydrogen bonds around the variant. In the case of p.Ser842Leu, there is a predicted reduction in the number of hydrogen bonds. Additionally, the site of hydrogen bond generation shifts from Tyr531 to Glu618 in the case of p.Arg583Trp.

### Clinical characteristics of the affected individuals

The patients with NOA displayed clinically significant reductions in testicular volume, while the patient diagnosed with cryptozoospermia exhibited normal testicular volume (Table 2). All three patients showed normal development of the penis, epididymis, vas deferens, and had no varicocele. Laboratory examinations indicated elevated levels of follicle-stimulating hormone (FSH) only in the NOA patients. The chromosomal karyotype of all three patients was 46,XY, without azoospermia factor microdeletions (Table 2). Semen analysis revealed that, while the semen volume in the patients was normal, sperm were absent in the NOA patients and occasionally present in the patient with cryptozoospermia (Table 2).

**Table 2.** Clinical characteristics of the NOA patients.

|  | Ref values | Patient 1<sup>b</sup> | Patient 2 | Patient 3 |
| --- | --- | --- | --- | --- |
| Age<sup>a</sup>(years) | - | 26-30 | 21-25 | 26-30 |
| Height/weight (cm/kg) | - | 179 /75 | 172/65 | 172/59 |
| Karyotype | 46,XY | 46,XY | 46,XY | 46,XY |
| Diagnosis | - | NOA | NOA | Cryptozoospermia |
| FSH (mIU/ml) | 0.95-11.95 | 14.31 | 22.37 | 2.4 |
| LH (mIU/ml) | 1.14-8.75 | 6.99 | 7.62 | 3.2 |
| PRL (ng/ml) | 3.46-19.40 | 9.44 | 9.9 | 6.73 |
| TSTO (ng/dl) | 187.17-950.28 | 299.369 | 387.499 | 410.000 |
| Semen volume (mL) | >1 . 4 | 2 | 4.2 | 2 |
| Sperm count (millions/mL) | >16 | 0 | 0 | 0 |
| Spermatozoa in centrifuged<br>semen sample<sup>c</sup> | - | 0 | 0 | Occasionally seen |
| Testis volume (mL) | >12 | L:10; R:NA | L:8; R:8 | L:13; R:12 |

Ref, reference; NOA, non-obstructive azoospermia; FSH, follicle-stimulating hormone; LH, luteinizing hormone; PRL, Prolactin; TSTO, Testosterone; L, left; R, right.
<sup>a</sup>Ages at the manuscript submission.
<sup>b</sup>F1: II-1 had previously undergone orchiectomy for cryptorchidism of the right testis.
<sup>c</sup>Centrifugation at 3000g for 15 minutes.

To investigate abnormalities in the spermatogenesis process, TESA was performed on P1 and P2. Testicular tissues from patient with obstructive azoospermia were used as a normal control (NC). Compaired to NC, HE staining of testicular tissue sections from the NOA patients with the *TBC1D8* variant revealed visible spermatogonia, spermatocytes, and round spermatids but lacked mature spermatozoa, indicating the phenotype of incomplete MA with impaired spermiogenesis rather than meiotic failure (Fig. 8a and Supplementary Fig. S3). TESA was initially performed to diagnose NOA. Subsequently, micro-TESE was carried out to retrieve sperm for ICSI. However, no viable spermatozoa were obtained by micro-TESE. Consequently, all NOA patients underwent in vitro fertilization using donor sperm and achieved successful pregnancy.

**Figure 8.**
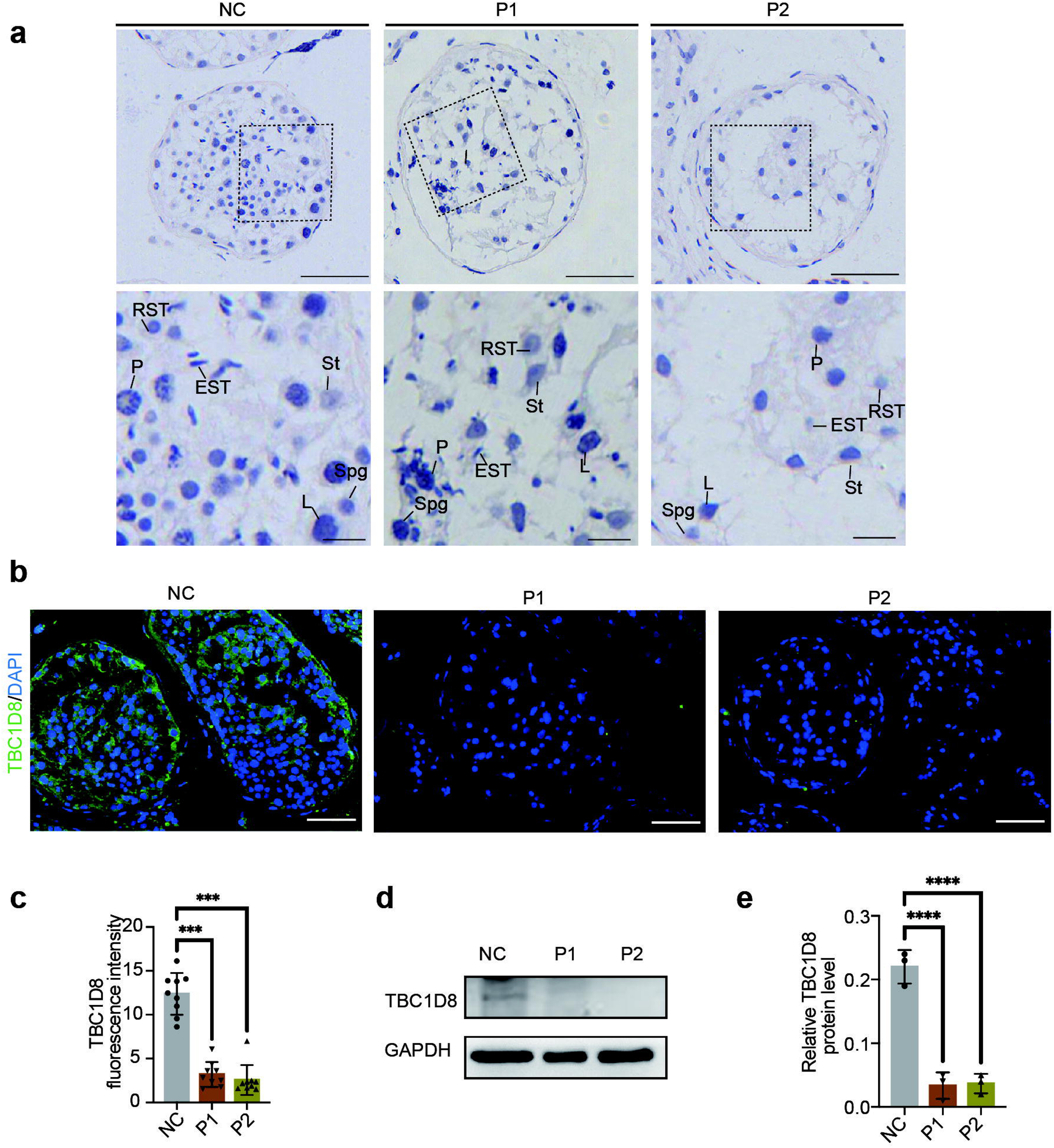
Pathogenic analysis of the candidate variants. (A) Hematoxylin staining analysis of testicular sections from Patient F1: II-1 (P1), Patient F2: II-1 (P2), and a normal control (NC) with obstructive azoospermia, illustrating the progression of spermatogenesis. Cross-sections of seminiferous tubules were acquired from testicular biopsy specimens. Spg, spermatogonia; L, leptotene spermatocyte; P, pachytene spermatocyte; RST, round spermatid; EST, elongating spermatids; St, Sertoli cells. Scale bars denote 50μm and 10μm. (B) Effects of the identified variants on TBC1D8 expression in the testes of NOA patients. TBC1D8 expression was assessed by immunofluorescence staining with antibodies against TBC1D8 (green) and DAPI (blue). Scale bars denote 50μm. (C) Fluorescence intensity of TBC1D8 in germ cells. Data are represented as individual values with mean ± SD. Unpaired 2-sided t test. (D) Protein expression of TBC1D8 in peripheral blood mononuclear cells from both the normal control and NOA patients. (E) Relative protein level of TBC1D8 in peripheral blood mononuclear cells after normalization to the GAPDH protein level. Data are represented as individual values with mean ± SD. The experiments were repeated three times. Unpaired 2-sided t test. \**P* < 0.05; \*\**P* < 0.01; \*\*\**P* < 0.001; \*\*\*\**P* < 0.0001.

### Effects of the identified variants on TBC1D8 expression

TBC1D8 is highly expressed at various stages of spermatogenesis in humans (Supplementary Fig. S4 a and b).To clarify the effect of the *TBC1D8* missense variant on protein expression, WB analysis of peripheral blood mononuclear cells, along with IF analysis of testicular tissue were conducted. TBC1D8 is mainly expressed in spermatogonia, spermatocytes, and spermatozoa in the control, but its absence in the NOA patients is consistent with the WB result (Fig. 8b-e). This suggests that the compound heterozygous missense variants may cause the decay of the TBC1D8 protein, resembling a likely knockout effect (Fig. 8d and e). These findings suggest that degradation of TBC1D8 may contribute to patients infertility.

## Discussion

The potential role of *TBC1D8* as a solitary genetic factor contributing to male infertility remains unexplored. For the first time, we report the presence of compound heterozygous variants of *TBC1D8* in sterile males revealing heterogeneity in clinical manifestations(21). variants in *TBC1D8* can lead to a decay in protein expression, resembling a knockout-like effect and causing severe spermatogenic dysfunction in affected individuals. *TBC1D*8 appears to play an essential role in male spermatogenesis, and a deficiency in TBC1D8 protein is likely to result in azoospermia, thereby impairing male fertility.

NOA significantly impairs male reproductive health. Elevated serum FSH levels and smaller testicular volumes are associated with more severe testicular histopathological patterns in men with NOA, indicating fewer remaining germ cells in the testis(22). Both P1 and P2 exhibited FSH levels exceeding the upper limit of the normal range, suggesting a decrease in spermatogenic function, consistent with incomplete MA observed in testicular sections. Notably, P2 showed a lower number of germ cells compared to P1, aligning with the difference in FSH levels and indicating more severe spermatogenic damage. In contrast, the patient with cryptozoospermia (pateint 3, P3) maintained sex hormones within the normal range. Individual sperm were occasionally found in P3 after semen centrifugation, while P1 and P2 do not. These results highlight the heterogeneity of pathogenicity caused by variants at different sites on the *TBC1D8* gene. This difference may reflect variable expressivity, as observed in other male infertility genes *TDRD9* (OMIM: 617963; incomplete MA and cryptozoospermia), *SYCP2* (OMIM: 604105; complete MA and cryptozoospermia), *TEX10* (OMIM: 616717; MA/SCOS, cryptozoospermia and oligozoospermia) and *TEX13A* (OMIM: 300312; SCOS and cryptozoospermia)(23–25). While occasional spermatozoa were observed in tissue sections from testicular punctures in NOA patients, no usable sperm could be obtained for ICSI treatment. Ultimately, three patients successfully conceived through the use of donor sperm. These observations suggest that TBC1D8 deficiency may be associated with not only a reduction in sperm quantity but also potential impairment of sperm quality in affected patients.

TBC1D8 comprises a TBC domain and two GRAM (gucosyl transferases RAB-like GTPase activators and myotubularins) domains, both exhibiting high conservation across eukaryotes. The TBC-containing RAB-specific GTPase-activating protein (TBC/RAB-GAP) facilitates the hydrolysis of RAB-GTP to RAB-GDP, promoting specific intracellular vesicle transport, pinocytosis, autophagy, cell division, and other essential biological processes(26). Loss-of- function variants in the *Tbc1d20* gene have been linked to azoospermia, while *Tbc1d21* gene variants are associated with teratozoospermia. Dysfunction in TBC/RAB-GAP leads to the accumulation of ubiquitinated autophagic cargo such as peroxisomes and mitochondria in testis tubules, resulting in defective sperm acrosome formation(6, 11). TBC/RAB-GAP also participates in intra-manchette transport during sperm tail formation, preventing tail rupture and reducing sperm motility(7, 27). Additionally, the TBC domain interacts with RAB proteins independently of its GAP activity, contributing to the formation and maintenance of the integrity of the mitochondrial sheath in the sperm tail(12). The GRAM domain potentially functions as a signaling domain, binding intracellular proteins or lipids and participating in membrane-related processes(28), promoting omegasome formation at the onset of autophagy by interacting with RAB protein(29). These findings underscore the significance of all domains in TBC1D8 functions. Moreover, variants occurring outside these domains may induce structural alterations, affecting the protein’s three-dimensional conformation, stability, and interactions with other molecules. In silico pathogenicity predictions vary across different variant sites, offering a potential explanation for the diverse phenotypes observed in TBC1D8 variants, resulting in both NOA and cryptozoospermia. In NOA patients, missense variants cause the decay of TBC1D8 protein. Data from *Tbc1d8^-/-^* mice testes replicated the NOA phenotype, supporting clinical observations. TBC1D8 interacts with multiple RAB proteins, with RAB8A scoring the highest. In the absence of TBC1D8 in the mice testes, the expression of RAB8A decreases, suggesting that RAB8A may function as an effector molecule of the TBC1D8 protein, rather than a substrate. TBC1D8 may depend on the interaction between the two domains and RAB8A positively regulating the expression of RAB8A. The catalytic effect of TBC1D8 on RAB8A needs further validation through additional experimentas.

In the process of spermatogenesis, autophagy plays a crucial role by mediating the degradation of damaged proteins and organelles through lysosomes, participating in acrosome biogenesis, flagella assembly, head shaping, and the removal of cytoplasm from elongating spermatids(11, 30). Autophagy-related genes are hub genes in spermatids, exhibiting stage- specific conseervation between humans and mice(31). Suppression and excess of autophagy were observed in the testes of patients with NOA patients, showing a strong association with impaired spermatogenesis(31, 32). At different stages of autophagy, distinct TBC proteins interact with specific RAB proteins, facilitating the coordination of membrane or vesicle trafficking(33, 34). In spermatozoa, RAB8A is localized in the acrosomal region, a specialized organelle derived from the Golgi apparatus(35, 36). RAB8A is involved in vesicular trafficking between the Golgi network and the plasma membrane, contributing to autophagy-based extracellular delivery(37, 38). RAB8A can interact with GM130 and regulate the distribution of the Golgi apparatus, playing a role in acrosome formation through the regulation of the autophagy pathway(39). Depletion of RAB8A accelerates the intracellular accumulation of LC3-II and p62, mediating the decreased extracellular secretion of Golgi-derived vesicles(40). Autophagic flux stalls, indicated by the accumulation of LC3 and P62, severely disturb acrosome formation(41). The current investigation reveals that TBC1D8 deficiency results in the accumulation of autophagosomes but fails in the fusion of autophagosomes with lysosomes due to a shortage of RAB8A. Moreover, while the increased expression of upstream ATG7 might partly alleviate the consequences of TBC1D8 deficiency and enhance autophagosome generation, it is unable to fully rectify the disruption in the fusion of autophagosomes and lysosomes resulting from RAB8A deficiency. This impediment hinders the elimination of residual bodies during spermiogenesis. The absence of RAB8A hinders the autophagic flux in *Tbc1d8^-/-^* mice, ultimately resulting in a shortage of spermatozoa with a mature acrosome, consistent with the phenotype observed in *Tbc1d20*- mutant mice. This study is the first to demonstrate that TBC1D8 positively regulates the expression of RAB8A and interacts with it to promote spermatozoa maturation through the autophagy pathway, specifically in acrosome formation and spermatozoa head shaping.

The crosstalk between apoptosis and autophagy is cruicial for testicular cell survival and death(42). Autophagy activation can mitigate spermatogenesis disorders induced by toxic substances through oxidative damage and inhibition of apoptosis (43). Lysosomal dysfunction and autophagy blockade contribute to cytotoxic death related to autophagy(44). Inhibiting autophagosome fusion with lysosomes notably reverses the downregulation of the BAX/BCL- 2 ratio and the expression of cleaved-Caspase3 (45). Autophagic flux blockade leads to insufficient substrates for the TCA cycle, inhibiting LDHA, and resulting in energy insufficiency, consequently leading to growth inhibition and apoptotic induction(46). The mitochondria-mediated intrinsic pathway is a major route for male germ cell apoptosis during spermatogenesis(47). Accumulation of pro-apoptotic Bax protein promotes the release of cytochrome c into the cytoplasm, initiating cell destruction via executioner-Caspase3 after Caspase-9 cleavage(48). The anti-apoptotic BCL2 protein act as a cellular guardian by inhibiting BAX activity and its translocation to the mitochondria(49). In male testes with MA, active Caspase-3 increases in the cytoplasm of germ cells, mediating the apoptotic process(50). RAB8A, reported as an oncogenic gene, promotes the proliferation, migration, and invasion of cancer cells(51). RAB8A knockdown significantly upregulates the expression of active Caspase-3 and BAX, while downregulating the expression of Caspase3 and BCL2 compared with the control group, ultimately promoting cell apoptosis(20, 52). Similarly, our findings indicate that apoptotic changes within the testes of *Tbc1d8^-/-^* mice predominantly involve elevated germ cell apoptosis, initiating the intrinsic mitochondrial pathway and exacerbating spermatocyte and spermatid loss. It is reasonable to infer that the increased germ cell apoptosis caused by TBC1D8 deficiency is mediated by RAB8A downregulation. Blockage of autophagic flux may further accelerate this apoptotic process via RAB8A in the testes of *Tbc1d8^-/-^*mice, resulting in azoospermia.

Previous studies have identified *TBC1D8* as an oncogene, demonstrating its involvement in tumorigenesis by upregulating genes associated with aerobic glycolysis and the cell cycle, such as cyclin D, GLUT, LDHA, c-myc, PDK1, and MEK5(13). Strong expression of TBC1D8 in the tumor microenvironment correlates with increased infiltration of immune cells, particularly M2 macrophages(14). In our study, three patients with *TBC1D8* variants did not develop any cancers. However, we recommend that patients carrying variants in *TBC1D8* undergo regular check-ups and screening for early detection, with timely intervention if cancer is identified.

Lastly, our study has some limitations. First, the number of clinical cases analyzed was limited, with only three patients carrying *TBC1D8* variants included in this study. Although all patients underwent microsurgical testicular sperm extraction, the small sample size and potential clinical heterogeneity restrict the generalizability of our findings. Therefore, conclusions regarding the impact of TBC1D8 deficiency on sperm quantity and quality should be interpreted with caution. Second, missense variants may have diverse functional consequences beyond simple loss-of-function, such as dominant-negative effects or gain-of- function, which cannot be fully modeled by a complete knockout. Thus, the full knockout mouse model used in this study may not fully recapitulate the molecular and phenotypic effects of patient-specific missense variants. Third, due to current technical and resource limitations, we have not yet established knock-in mouse models carrying the specific missense mutations, nor conducted in vitro assays (e.g., protein stability, cellular localization, or rescue experiments) to directly validate the functional impact of these variants. Furthermore, we have not fully characterized how these variants affect downstream molecular pathways, such as those related to autophagy, apoptosis, or germ cell development, which are implicated in our mouse model. These mechanistic studies will be crucial for delineating the pathophysiological relevance of *TBC1D8* variants. Finally, although our findings suggest a potential role for TBC1D8 in male fertility, further investigations involving larger patient cohorts, variant-specific functional studies, and pathway-level analyses are necessary to substantiate the proposed pathogenic mechanisms and their clinical implications.

Collectively, our findings strongly suggest that variants in *TBC1D8* likely underlie spermatogenic failure in a subgroup of men with azoospermia. TBC1D8 deficiency disrupts autophagic flux and enhances apoptosis by losing its coupling with RAB8A, ultimately resulting in azoospermia and infertility. Given the evidence that TBC1D8 deficiency is associated with impaired spermiogenesis and NOA, we propose that genetic testing for *TBC1D8* may serve as a valuable and cost-effective diagnostic tool in male infertility clinics. In particular, it may be considered in NOA patients with idiopathic etiology, especially when standard hormonal, cytogenetic, and AZF microdeletion analyses yield normal results. Although *TBC1D8* variants are rare, incorporating *TBC1D8* into targeted gene panels or exome sequencing workflows, followed by Sanger sequencing for validation, offers a cost- effective model for genetic diagnosis. Early identification of pathogenic variants not only informs prognosis and guides individualized treatment decisions but also facilitates genetic counseling for affected patients and their families. As sequencing technologies continue to become more accessible and affordable, integrating *TBC1D8* testing into routine diagnostic pipelines for idiopathic NOA is both clinically relevant and economically feasible.

## Data Availability

All data produced in the present work are contained in the manuscript

## Acknowledgements

We thank all participants and their families, as well as the groups and physicians who supported this study. We thank all the doctors, nurses, and embryologists in the Reproductive Medicine Center of the Second Xiangya Hospital for their clinical work.

## Authors contributions

Co-first authorship order was decided by weighing productivity and significance of work. Conceptualization: H.Z, Zeng.J, H.H; Data curation: H.Z; Formal analysis: H.F, Y.R, G.J, J.K, J.J; Funding acquisition: H.H; Investigation: Zeng.J; Methodology: H.Z, Zeng.J, H.H, Zhang.J, L.Z; Project administration: X.H, C.J,and H.H.; Resources: C.J,and H.H.; Supervision: X.H, C.J, H.H; Validation: H.Z, Zeng.J, Q.R, Zhang.J, X.R, L.Z, W.M; Visualization: Q.R, X.R, W.M; Writing-original draft: H.Z, Zeng.J; Writing-review & editing: Zhang.J, Zhao.J, Y.K, X.H, X.H, C.J,H.H, Zhao.J.

## Ethics declaration

The animal study was reviewed and approved by the Animal Ethics Committee of Hunan University of Traditional Chinese Medicine. All methods were performed in accordance with the relevant guidelines and regulations. For study patients, this study was approved by the Ethics Committee of the Second Xiangya Hospital, Central South University. Written informed consent was obtained from all human participants.

## Supplementary Figure Legends

**Figure S1.**
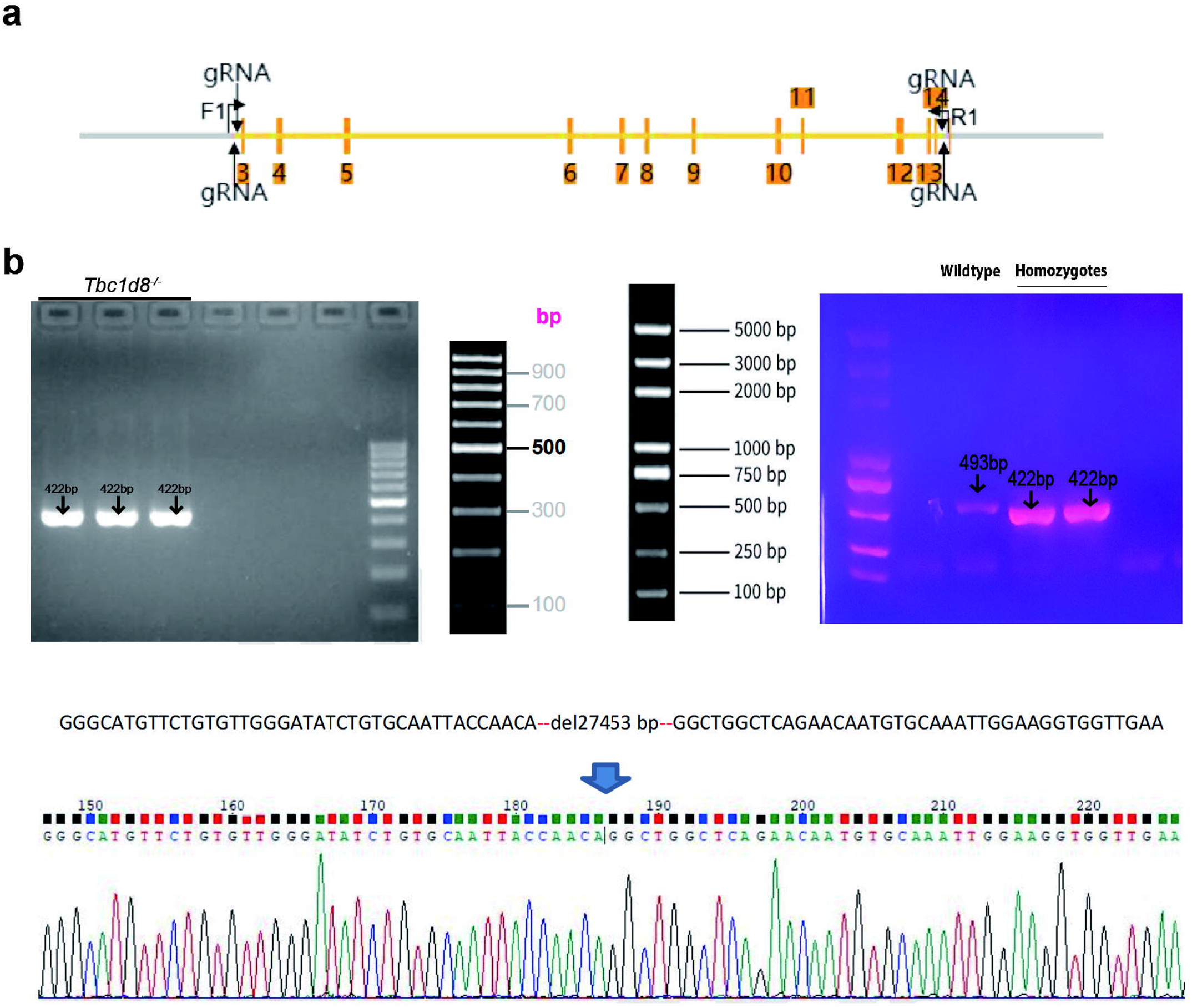
Generation and Validation of *Tbc1d8* Knockout Mice. (A) Schematic diagram of *Tbc1d8* knockout. CRISPR/Cas9 technology was used to generate Tbc1d8 knockout (*Tbc1d8^-/-^*) mice by deleting exon 3 to exon 14. (B) Sequencing Confirmation of knockout effect in *Tbc1d8^-/-^* mice. Homozygotes: one band with 422 bp. Heterozygotes: two bands with 422 bp and 493 bp. Wild-type allele: one band with 493 bp.

**Figure S2.**
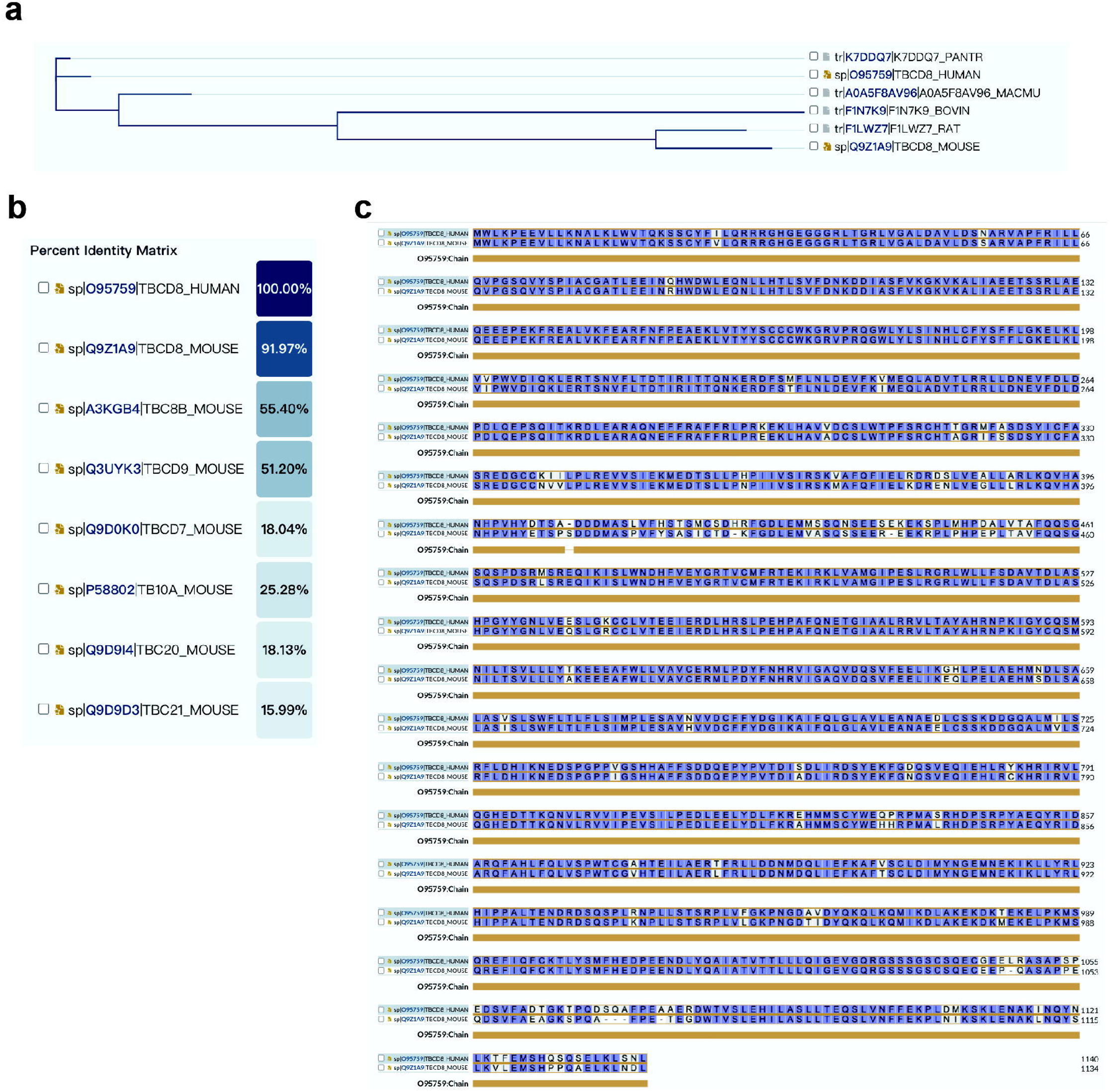
Sequence alignment of TBC1D8 from different species. (A) Phylogenetic tree of TBC1D8 protein from different species. (B) Alignment of amino acids of TBC1D8 from human and TBC proteins from mice. (C) Alignment of amino acids of TBC1D8 from human and mice.

**Figure S3.**
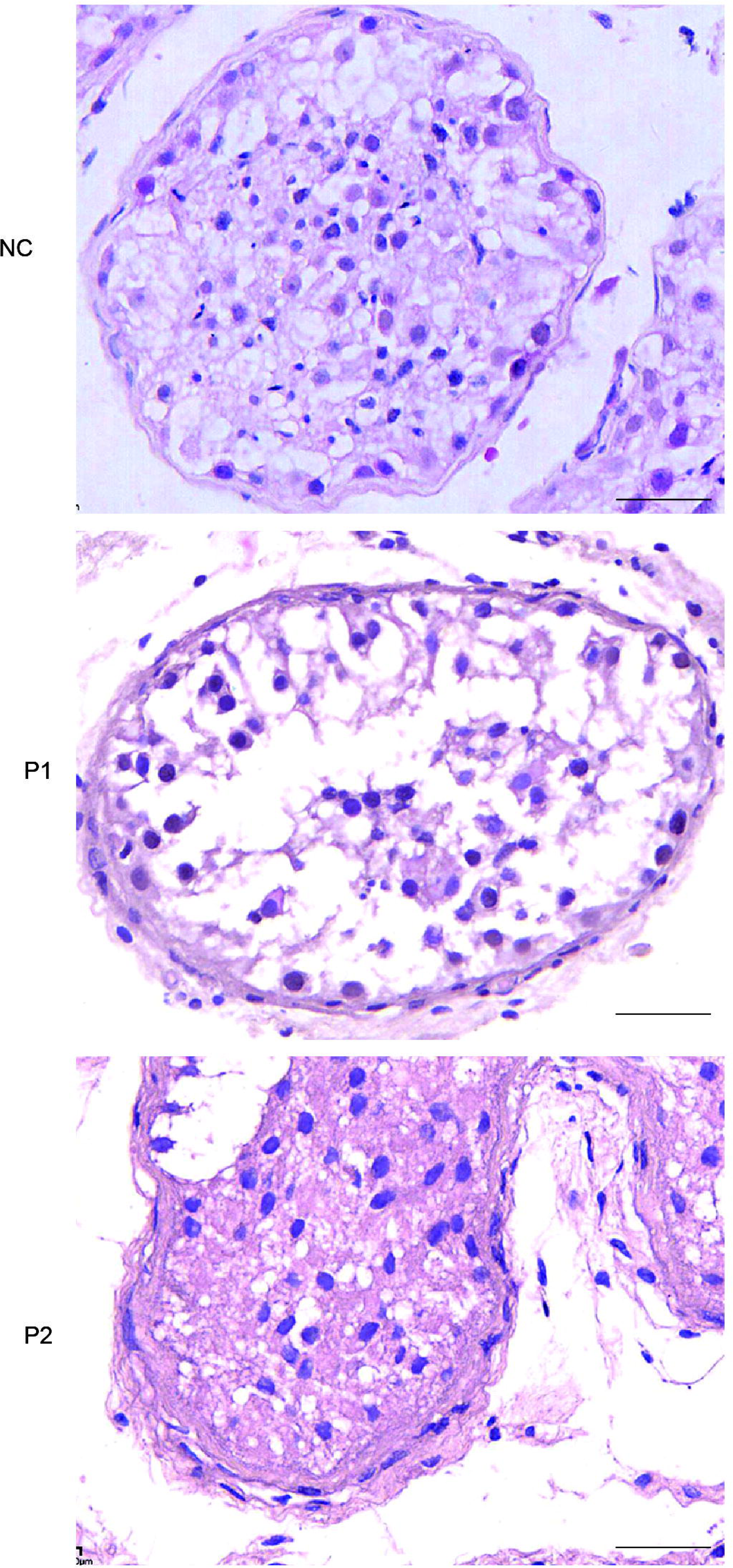
Clinical phenotypes. H&E staining of cross sections of seminiferous tubules using testicular biopsy samples from patient F1: II-1(P1), patient F2: II-1(P2), and a normal control (NC) with obstructive azoospermia, illustrating the progression of spermatogenesis. Scale bars denote 25μm.

**Figure S4.**
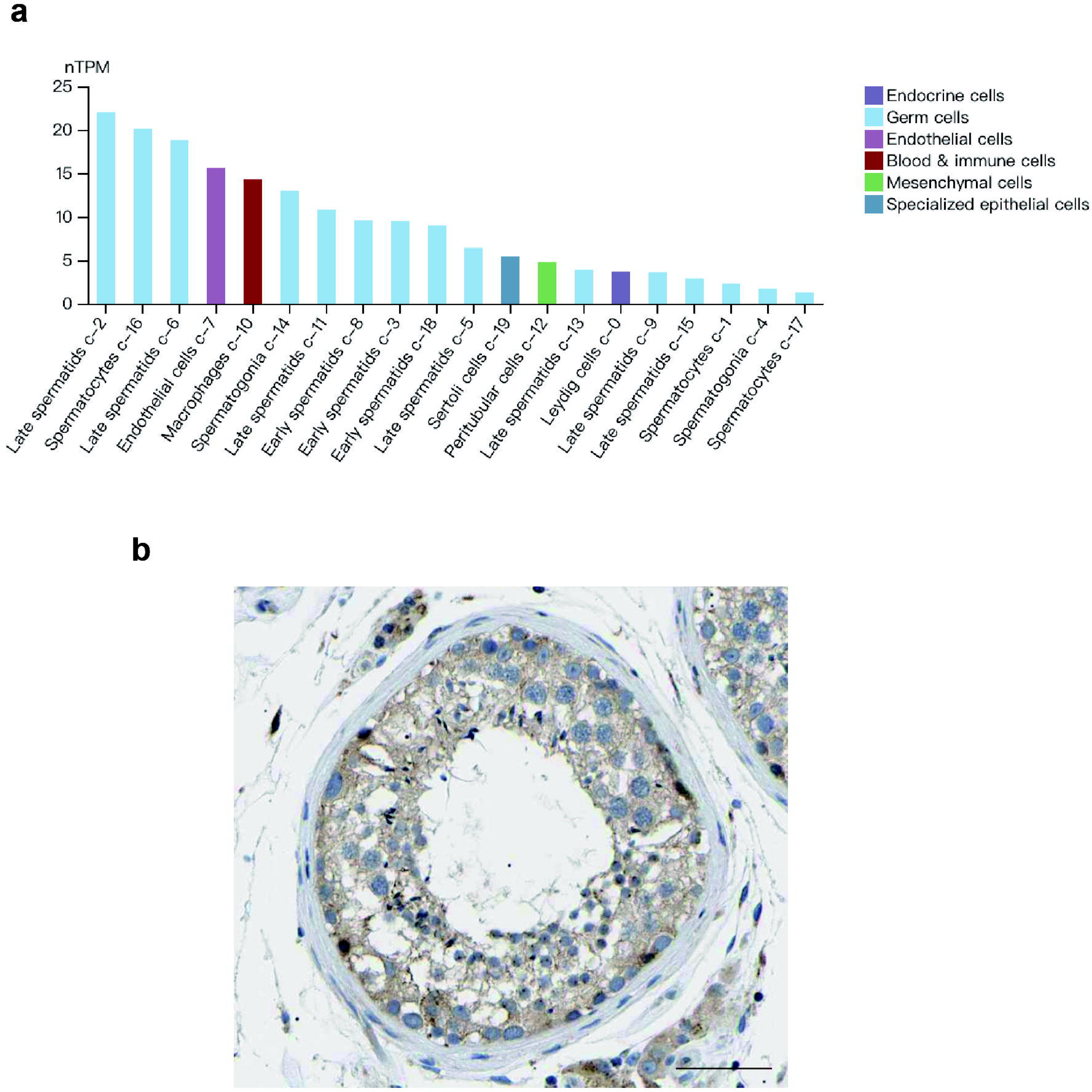
Expression pattern of *TBC1D8* from HUMAN PROTEIN ATLAS database. (A) Analysis of *TBC1D8* mRNA expression in normal human testes using single-cell transcriptomic data. The data reveal the spatial and cellular distribution of *TBC1D8* expression within the testicular tissue, providing insights into its specific localization. (B) Immunohistochemical analysis of TBC1D8 signals in a normal testicular section obtained from a healthy middle-aged man. The staining pattern highlights TBC1D8 localization in specific cell types within the testicular tissue. Scale bars denote 25μm.

## Supplementary Tables

**Table S1.** *TBC1D8* variants identified in the NOA patients.

| TBC1D8 variants | MT1 | MT2 | MT3 | MT4 | MT5 |
| --- | --- | --- | --- | --- | --- |
| cDNA alteration | c.3322A>G | c.1747C>T | c.2566A>G | c.845C>T | c.2525C>T |
| Protein alteration | p. Met1108Val | p. Arg583Trp | p. Ile856Val | p. Ala282Val | p. Ser842Leu |
| Variant type | Missense | Missense | Missense | Missense | Missense |
| SIFT <sup>a</sup> | Tolerable | Damaging | Tolerable | Damaging | Tolerable |
| Mutation Taster <sup>b</sup> | Disease causing | Disease causing | Polymorphism | Disease causing | Polymorphism |
| PROVEAN <sup>c</sup> | Tolerable | Damaging | Tolerable | Damaging | Tolerable |
| CADD <sup>d</sup> | Tolerable | Damaging | Tolerable | Damaging | Tolerable |
| FATHMM_MKL <sup>e</sup> | Damaging | Damaging | Damaging | Damaging | Tolerable |
| All individuals in<br>gnomAD <sup>f</sup> | 0.003171 | 0.001003 | 0.000054 | 0.000016 | 0.000083 |
| ExAC ALL <sup>g</sup> | 0.009614 | 0.000145 | 0.000313 | 0.001341 | 0.000109 |

NA, not available; MT1–5, mutations 1–5.
<sup>a</sup>SIFT, Sorting Intolerant from Tolerant https://sift.bii.a-star.edu.sg/.
<sup>b</sup>Mutation Taster, https://www.mutationtaster.org/.
<sup>c</sup>PROVEAN, Protein Variation Effect Analyzer https://www.jcvi.org/research/provean.
<sup>d</sup>CADD, Combined Annotation Dependent Depletion https://cadd.gs.washington.edu/.
<sup>e</sup>FATHMM\_MKL, Functional Analysis through Hidden Markov Models-Multiple Kernel Learning https://fathmm.biocompute.org.uk/fathmmMKL.htm.
<sup>f</sup>gnomAD, Genome Aggregation Database https://gnomad.broadinstitute.org/.
<sup>g</sup>ExAC, the Exome Aggregation Consortium, https://gnomad.broadinstitute.org/.

**Table S2.**
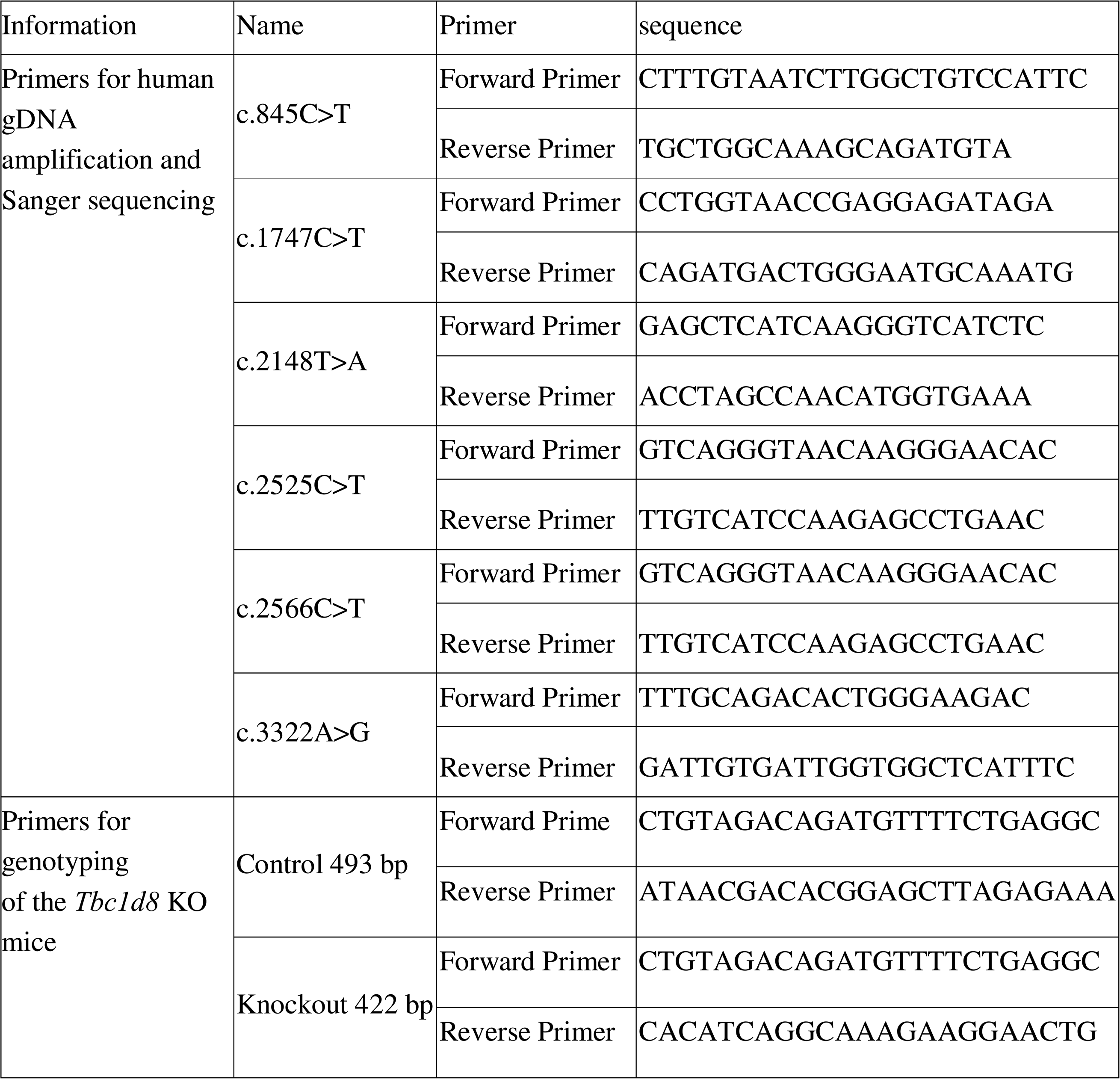
Primers used in this study.

**Table S3.**
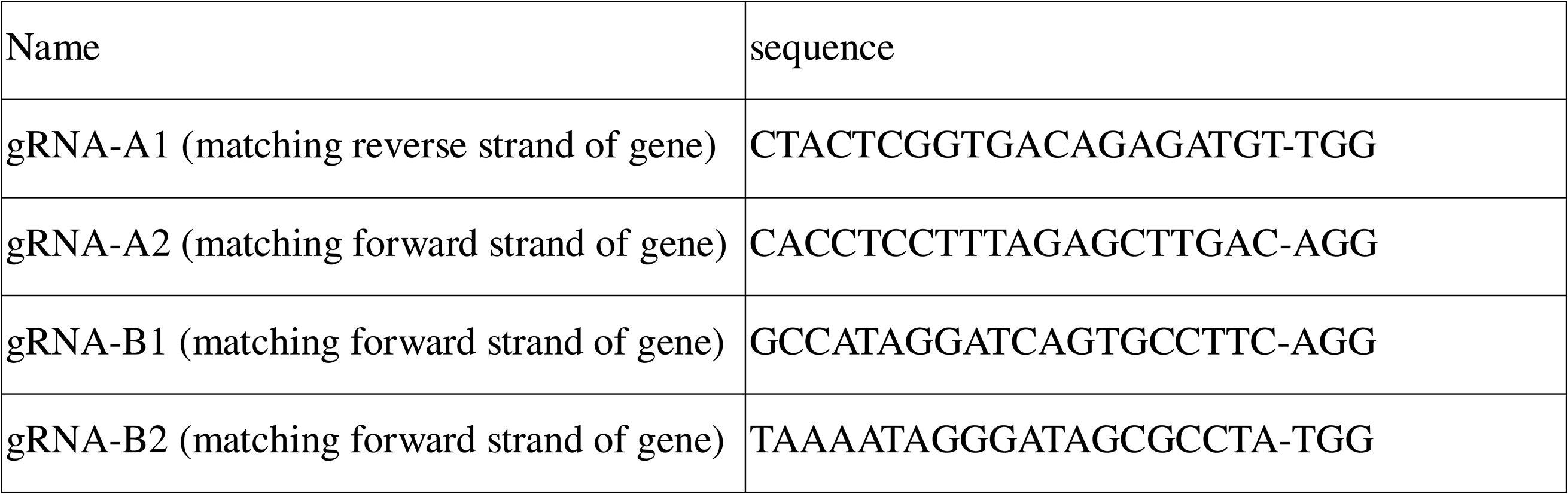
SgRNA used for generation of *Tbc1d8* KO mice

**Table S4.** Antibodies used in this study.

| Protein name | Catalogue number | Manufactur<br>e | Application |
| --- | --- | --- | --- |
| TBC1D8 | sc-100872 | Santacruz | WB (1:1000)<br>IF(1:100) |
| ATG7 | Ab52472 | Abcam | IF(1:200)<br>WB: (1:10000) |
| ATG5 | R381320 | Zenbio | WB (1:1000) |
| ATG3 | R23552 | Zenbio | WB (1:1000) |
| LC3 | 14600-1-AP | Proteintech | WB (1:1000)<br>IF(1:200) |
| RAB8A | 67536-1-Ig | Proteintech | WB (1:1000)<br>IF(1:200) |
| DDX4 | 51042-1-AP | Proteintech | WB (1:1000)<br>IF(1:200) |
| BAX | R22708 | Zenbio | WB (1:1000)<br>IF(1:200) |
| BCL2 | R23309 | Zenbio | WB (1:1000)<br>IF(1:25) |
| Cleaved-caspase3 | 300968 | Zenbio | WB (1:1000)<br>IF(1:100) |
| P62 | R25788 | Zenbio | WB (1:1000)<br>IF(1:200) |
| GAPDH-HRP | HRP-60004 | Proteintech | WB (1:10000) |
| α-tubulin | 66031-1-Ig | Proteintech | WB (1:1000) |
| Alexa Fluor 594-conjugated<br>Donkey Anti-rabbit IgG | SA00013-4 | Proteintech | IF(1:200) |
| Alexa Fluor 488-conjugated<br>Donkey Anti-mouse IgG | SA00013-1 | Proteintech | IF(1:200) |
| HRP-conjugated Donkey<br>Anti-rabbit IgG | HA1001 | HUABIO | WB ( 1:5000 ) |

